# Psychotropic drugs in Portugal from 2016 to 2019: a nationwide pharmacoepidemiological profile

**DOI:** 10.1101/2022.09.14.22279819

**Authors:** Luís Madeira, Guilherme Queiroz, Rui Henriques

## Abstract

**Background:** The prescription of psychotropic medication is rising in Europe along the last decade. Exploring consumption patterns in pre-pandemic times in Portugal, as well as relevant socio-demographic determinants, can help establish comparisons with worldwide patterns and support public health policies for mental health.

**Methods:** Descriptive, non-comparative cohort study, comprising full nationwide drug prescription records in Portugal along antidepressant, antipsychotic, and anxiolytic classes. Statistical analysis of prescription and consumption patterns according to reference dosages and guided by several criteria, including active substance, demographics, geography, associated medical specialty, and incurred costs.

**Results:** An increase of 29.6% and 34.7% in the consumption of antipsychotics and antidepressants between 2016 and 2019 is highlighted, reasonably accompanied by an increase of 37M Eur in total expenditure (>20M Eur in public copay) for these classes of drugs. Disparities in sociodemographic and geographical incidence are identified. Amongst other pivotal results, we further observed that 64% of psychotropic drug prescriptions are undertaken by general practitioners, while only 21% undertaken by neurological and psychiatric specialties.

**Conclusion:** Nationwide patterns of psychotropic drug prescription in Portugal reveal notable trends and determinants, establishing a reference point for cross-regional studies and being currently assessed at a national level to establish psychosocial initiatives and guidelines for the clinical practice and medical training.

**Novelty:** To our knowledge, first Portuguese psychopharmacoepidemiological study assessing: 1) economic correlates; 2) prescription patterns by medical specialty; 3) adherence rates and geographical determinants; 4) consumption patterns by active substance; and 5) systematic trends for the pre-pandemic period.

## 1 Introduction

Psychiatry disorders are among the leading chronic non-communicable diseases in the world (IHME 2022), and Portugal is no exception, with a prevalence of 18.4% of mental illness. The commitment to deinstitutionalization and the development of community-based mental health services has fortunately widened the access to psychotropic drugs and represents a step forward in mental health care. The consumption of psychotropic drugs, namely antidepressants, antipsychotics and anxiolytics, reported increasing trends in the last two decades both worldwide (OECD 2020; Bachhuber *et al*. 2016; Murphy *et al*. 2016; Xu *et al*. 2021; Hálfdánarson *et al*. 2017) and in Portugal (Caldas de Almeida *et al*. 2013), especially among women and the elderly (Semoun and Sevilla-Dedieu 2015; Carrasco-Garrido *et al*. 2016). However, thorough surveillance is important to tackle issues concerning inequalities in access, overuse of psychotropics like benzodiazepines, and inadequate active drug selection (Gardner 2014).

Portugal is a paradigmatic case of heavy use of anxiolytics since the 90s (Saúde 2019), as this class represents 2% of all drugs sold and the country holds the higher consumption rate among OECD countries. However, the prescription trend of this therapeutic group has remained stable (OECD and EU 2018). As for antidepressants, Portugal is the fifth OECD country with highest consumption rate, with past data evidencing increasing trends similar to other countries, like the UK, where it almost doubled in the last decade (Saúde 2019). Finally, antipsychotic consumption has also registered considerable upward trends, even though Portugal kept an average consumption below the OECD average (Saúde 2019). However, most data on consumption is outdated, especially concerning antipsychotics. Besides, by focusing only on consumption rates, it fails to comprehend the dynamics of access to mental healthcare, the speciality responsible for that care and medication adherence. This information is vital to health policy and planning, and can help to design more effective and tactical campaigns aiming to improve the quality of prescription, the communication between primary and hospital care, and non-pharmacological support.

Most health data in Portugal is currently fully digital and centralized in the Shared Services Ministry of Health (SPMS), comprehending data on demographics and clinical records, and an electronic prescription platform (PEM) that allows tracing of every prescription, both public and private. This represents an excellent opportunity in the European context to study nationwide pharmacoepidemiological factors and offer a systematic assessment and characterization of the prescription and consumption trends of psychiatric drugs from 2016 to 2019.

The proposed epidemiological study is the first one comprehensively traversing the Portuguese status on psychotropic drug prescription along the years preceding the COVID 19 pandemic. In addition, it further reveals sociodemographic determinants, the incidence of prescriptions per medical specialty, the associated expenditures, geographic discrepancies, and compliance rates.

## 2 Methods

We performed a descriptive non-comparative cohort study, with data related to all users in Portugal with registered prescriptions of any antipsychotic, antidepressant, or benzodiazepine approved for commercial usage by Infarmed (active principles listed in Appendix B1) from 1/1/2016 to 31/12/2019. For each user (granted anonymity), we collected data on gender, age group (10-year ranges), residency, and their respective prescriptions. For each prescription, we collected the medical prescription identifier, the medical speciality of the prescriber, the week of prescription, the municipality where it took place, and a package code with information on active principle, commercial name, number of units, dosage form and pharmaceutical formulation.

All data was mapped onto a relational database where the profile of the user, geographical aspects, the description of the drugs, and the characteristics of the packages in the prescriptions were decoupled to promote time and memory efficiency associated with data exploration tasks. Data were preprocessed to correctly map equivalent coding.

The number of residents per municipality and national district for different gender and age groups were collected from the portal Instituto Nacional de Estatística (INE).

The users’ municipality of residence was inferred from their primary health care centre, not from the place of prescription, which is often carried out in central hospitals.

Drug prescription is expressed in Defined Daily Dose per 1000 inhabitants-days (DID). The Defined Daily Dose (DDD) corresponds to the assumed average maintenance dose per day for a drug used for its main indication in adults. For this, we first assigned a DDD to each active ingredient in the study according to its ATC code, using the guidelines of the World Health Organization (WHO). In the case of drugs without an ACT code, a DDD value was assigned based on previous studies with the same active ingredient^1^. The DDD units per package were calculated and assigned to their identifier. The complete list of packages and correspondent DDD can be consulted in Appendix B2. DID of each active ingredient was then obtained by multiplying the DDD units per package by its total prescription and divided by the target population size and period of study.

To account for meaningful deviations, the geographic distribution of DID statistics are standardized by the age distribution of citizens along the Portuguese territory, unless stated otherwise.

The data processing, user location estimates, and subsequent analytical tasks were conducted using Python and PostgreSQL. The graphical presentation of statistics was carried out in Plotly.

## 3 Results

### Birds-eye view of data

This study covers a total of 46,161,485 prescriptions, of which 17,529,112 correspond to antidepressants, 6,541,283 to antipsychotics, and 22,091,090 to benzodiazepines. Tables 1-4 summarize the yearly statistics of the national cohort across drug classes, gender, age, and geography. Table 1 presents the distribution of consumption rates in DIDs; Table 2 assesses the adherence rate (consumption to prescription DID ratio); Table 3 provides the number of prescribed users; and Table 4 summarizes the total expenditures.

**Table 1.** Summary of the psychodrug consumption status in Portugal: consumption rates across demographic variables expressed in Defined Daily Dose per 1000 inhabitants-days (DIDs).

|  |  | antipsychotics |  |  |  | antidepressants |  |  |  | benzodiazepines |  |  |  |
| --- | --- | --- | --- | --- | --- | --- | --- | --- | --- | --- | --- | --- | --- |
|  |  | 2016 | 2017 | 2018 | 2019 | 2016 | 2017 | 2018 | 2019 | 2016 | 2017 | 2018 | 2019 |
| consumption rates (DIDs) | all | 10.93 | 12.90 | 13.38 | 14.17 | 83.03 | 97.78 | 103.66 | 111.82 | 57.86 | 65.31 | 65.00 | 64.33 |
|  | gender |  |  |  |  |  |  |  |  |  |  |  |  |
|  | F | 10.18 | 12.06 | 12.59 | 13.31 | 121.43 | 142.30 | 150.37 | 161.69 | 77.22 | 86.91 | 86.14 | 85.00 |
|  | M | 11.76 | 13.84 | 14.24 | 15.11 | 40.65 | 48.63 | 52.10 | 56.78 | 36.49 | 41.46 | 41.65 | 41.52 |
|  | age |  |  |  |  |  |  |  |  |  |  |  |  |
|  | [18-29] | 5.02 | 5.89 | 6.23 | 6.75 | 17.24 | 20.25 | 21.88 | 24.76 | 6.20 | 6.80 | 6.68 | 6.69 |
|  | [30-39] | 9.37 | 10.49 | 10.26 | 10.52 | 42.02 | 45.94 | 45.91 | 47.46 | 20.91 | 21.86 | 20.59 | 19.56 |
|  | [40-49] | 14.65 | 16.94 | 17.10 | 17.84 | 81.68 | 94.18 | 97.93 | 103.97 | 49.08 | 54.10 | 52.97 | 51.80 |
|  | [50-59] | 16.68 | 19.69 | 20.26 | 21.45 | 118.94 | 137.33 | 142.78 | 151.99 | 81.79 | 91.79 | 90.83 | 89.13 |
|  | [60-69] | 15.08 | 18.06 | 18.93 | 20.24 | 154.83 | 181.15 | 191.92 | 205.38 | 114.65 | 129.98 | 129.26 | 127.30 |
|  | [70-79] | 14.53 | 17.41 | 18.62 | 19.80 | 184.41 | 220.94 | 237.61 | 259.98 | 140.91 | 159.64 | 160.08 | 159.77 |
|  | [+80] | 22.25 | 27.88 | 30.35 | 32.86 | 195.68 | 248.02 | 275.85 | 306.71 | 162.92 | 191.37 | 195.63 | 198.28 |
|  | region |  |  |  |  |  |  |  |  |  |  |  |  |
|  | alentejo | 10.83 | 13.18 | 13.82 | 14.63 | 78.33 | 91.22 | 96.81 | 102.85 | 45.26 | 50.06 | 48.80 | 47.92 |
|  | algarve | 10.08 | 11.54 | 12.40 | 13.62 | 50.54 | 59.59 | 64.21 | 70.13 | 35.11 | 40.42 | 41.25 | 42.33 |
|  | centro | 12.10 | 13.96 | 14.50 | 15.26 | 87.59 | 100.60 | 105.89 | 113.70 | 62.56 | 70.01 | 69.76 | 68.56 |
|  | lisboa | 11.54 | 14.02 | 14.58 | 15.29 | 79.77 | 96.10 | 102.28 | 110.61 | 42.86 | 49.11 | 48.66 | 48.42 |
|  | norte | 9.92 | 11.58 | 11.89 | 12.73 | 87.42 | 103.27 | 109.64 | 118.78 | 72.01 | 81.30 | 81.22 | 80.49 |

**Table 2.** Therapeutic adherence in Portugal: consumption to prescription DIDs ratio per demographic group.

|  |  | antipsychotics |  |  |  | antidepressants |  |  |  | benzodiazepines |  |  |  |
| --- | --- | --- | --- | --- | --- | --- | --- | --- | --- | --- | --- | --- | --- |
|  |  | 2016 | 2017 | 2018 | 2019 | 2016 | 2017 | 2018 | 2019 | 2016 | 2017 | 2018 | 2019 |
| adherence ratio | all | 0.88 | 0.82 | 0.81 | 0.81 | 0.87 | 0.79 | 0.79 | 0.79 | 0.97 | 0.96 | 0.96 | 0.96 |
|  | gender |  |  |  |  |  |  |  |  |  |  |  |  |
|  | F | 0.87 | 0.80 | 0.80 | 0.80 | 0.86 | 0.79 | 0.79 | 0.79 | 0.97 | 0.96 | 0.96 | 0.96 |
|  | M | 0.88 | 0.83 | 0.82 | 0.82 | 0.87 | 0.80 | 0.79 | 0.79 | 0.97 | 0.97 | 0.96 | 0.96 |
|  | age |  |  |  |  |  |  |  |  |  |  |  |  |
|  | [18-29] | 0.86 | 0.80 | 0.79 | 0.78 | 0.86 | 0.77 | 0.76 | 0.76 | 0.96 | 0.95 | 0.95 | 0.94 |
|  | [30-39] | 0.87 | 0.81 | 0.80 | 0.80 | 0.86 | 0.77 | 0.77 | 0.77 | 0.97 | 0.95 | 0.95 | 0.95 |
|  | [40-49] | 0.87 | 0.82 | 0.81 | 0.80 | 0.86 | 0.78 | 0.78 | 0.78 | 0.97 | 0.95 | 0.95 | 0.95 |
|  | [50-59] | 0.87 | 0.81 | 0.81 | 0.80 | 0.85 | 0.78 | 0.78 | 0.78 | 0.97 | 0.96 | 0.96 | 0.95 |
|  | [60-69] | 0.88 | 0.82 | 0.81 | 0.81 | 0.87 | 0.80 | 0.80 | 0.80 | 0.97 | 0.96 | 0.96 | 0.96 |
|  | [70-79] | 0.88 | 0.82 | 0.82 | 0.81 | 0.87 | 0.81 | 0.80 | 0.80 | 0.98 | 0.97 | 0.97 | 0.97 |
|  | [+80] | 0.90 | 0.83 | 0.83 | 0.82 | 0.89 | 0.82 | 0.81 | 0.81 | 0.98 | 0.97 | 0.96 | 0.96 |
|  | region |  |  |  |  |  |  |  |  |  |  |  |  |
|  | alentejo | 0.85 | 0.79 | 0.77 | 0.78 | 0.84 | 0.78 | 0.77 | 0.77 | 0.96 | 0.95 | 0.95 | 0.95 |
|  | algarve | 0.87 | 0.78 | 0.78 | 0.77 | 0.85 | 0.74 | 0.75 | 0.75 | 0.97 | 0.95 | 0.95 | 0.95 |
|  | centro | 0.87 | 0.82 | 0.82 | 0.81 | 0.86 | 0.79 | 0.80 | 0.80 | 0.97 | 0.96 | 0.96 | 0.96 |
|  | lisboa | 0.87 | 0.80 | 0.79 | 0.79 | 0.86 | 0.78 | 0.77 | 0.78 | 0.97 | 0.96 | 0.96 | 0.95 |
|  | norte | 0.89 | 0.84 | 0.84 | 0.83 | 0.88 | 0.81 | 0.81 | 0.81 | 0.97 | 0.97 | 0.97 | 0.96 |

**Table 3.** Number of prescribed individuals with psychotropic drugs per demographic group.

|  |  | antipsychotics |  |  |  | antidepressants |  |  |  | benzodiazepines |  |  |  |
| --- | --- | --- | --- | --- | --- | --- | --- | --- | --- | --- | --- | --- | --- |
|  |  | 2016 | 2017 | 2018 | 2019 | 2016 | 2017 | 2018 | 2019 | 2016 | 2017 | 2018 | 2019 |
| number of users | all | 388,305 | 417,156 | 432,809 | 446,390 | 1221,550 | 1305,811 | 1358,035 | 1421,671 | 1506,424 | 1561,381 | 1537,151 | 1523,839 |
|  | gender |  |  |  |  |  |  |  |  |  |  |  |  |
|  | F | 238,947 | 256,735 | 266,587 | 274,631 | 911,818 | 969,431 | 1004,136 | 1046,860 | 1041,085 | 1073,270 | 1053,174 | 1041,837 |
|  | M | 149,358 | 160,421 | 166,222 | 171,759 | 309,732 | 336,380 | 353,899 | 374,811 | 465,339 | 488,111 | 483,977 | 482,002 |
|  | age |  |  |  |  |  |  |  |  |  |  |  |  |
|  | [18-29] | 19,853 | 21,398 | 22,186 | 23,546 | 54,576 | 59,380 | 61,886 | 67,373 | 54,585 | 57,737 | 58,612 | 59,670 |
|  | [30-39] | 32,060 | 32,222 | 31,393 | 31,175 | 107,118 | 107,940 | 106,091 | 107,395 | 110,312 | 110,130 | 104,147 | 101,252 |
|  | [40-49] | 54,747 | 57,949 | 58,791 | 59,257 | 191,059 | 202,614 | 207,224 | 214,524 | 205,365 | 213,349 | 208,174 | 205,690 |
|  | [50-59] | 64,775 | 69,473 | 71,299 | 72,917 | 237,559 | 251,211 | 258,342 | 267,456 | 277,900 | 287,245 | 279,901 | 275,419 |
|  | [60-69] | 63,728 | 68,926 | 71,336 | 73,042 | 246,970 | 263,974 | 275,104 | 286,604 | 321,406 | 333,187 | 326,665 | 321,077 |
|  | [70-79] | 65,931 | 70,339 | 73,267 | 75,663 | 220,607 | 237,516 | 251,242 | 265,031 | 302,742 | 311,900 | 309,532 | 308,738 |
|  | 80+ | 87,211 | 96,849 | 104,537 | 110,790 | 163,661 | 183,176 | 198,146 | 213,288 | 234,114 | 247,833 | 250,120 | 251,993 |

**Table 4.** Total expenditure (kEur) and governmental participation (kEur) with psychotropic drugs in Portugal.

|  |  |  | antipsychotics |  |  |  | antidepressants |  |  |  | benzodiazepines |  |  |  |
| --- | --- | --- | --- | --- | --- | --- | --- | --- | --- | --- | --- | --- | --- | --- |
|  |  |  | 2016 | 2017 | 2018 | 2019 | 2016 | 2017 | 2018 | 2019 | 2016 | 2017 | 2018 | 2019 |
| volume of sales (1000Eur) | all |  | 54,925 | 59,728 | 60,414 | 63,290 | 23,908 | 28,254 | 29,810 | 32,355 | 14,671 | 16,040 | 15,771 | 15,655 |
|  | region | alentejo | 4,465 | 4,915 | 4,989 | 5,218 | 1,983 | 2,325 | 2,461 | 2,658 | 1,097 | 1,180 | 1,140 | 1,112 |
|  |  | algarve | 2,089 | 2,305 | 2,597 | 2,850 | 710 | 818 | 863 | 946 | 463 | 513 | 518 | 530 |
|  |  | centro | 14,330 | 15,437 | 15,883 | 16,691 | 6,345 | 7,402 | 7,730 | 8,295 | 3,848 | 4,226 | 4,160 | 4,124 |
|  |  | lisboa | 16,985 | 19,098 | 19,564 | 20,211 | 6,046 | 7,281 | 7,709 | 8,387 | 3,172 | 3,509 | 3,429 | 3,396 |
|  |  | norte | 17,056 | 17,974 | 17,381 | 18,320 | 8,825 | 10,428 | 11,047 | 12,069 | 6,090 | 6,612 | 6,525 | 6,492 |
| public copay (1000Eur) | all |  | 62,828 | 71,128 | 72,268 | 74,845 | 66,464 | 77,535 | 81,878 | 88,827 | 32,878 | 36,868 | 36,859 | 36,873 |
|  | region | alentejo | 5,081 | 5,850 | 5,949 | 6,136 | 5,181 | 5,988 | 6,325 | 6,782 | 2,377 | 2,618 | 2,573 | 2,544 |
|  |  | algarve | 2,390 | 2,728 | 3,077 | 3,330 | 2,084 | 2,393 | 2,525 | 2,741 | 1,092 | 1,244 | 1,281 | 1,321 |
|  |  | centro | 16,404 | 18,377 | 18,915 | 19,631 | 17,164 | 19,477 | 20,343 | 21,859 | 8,447 | 9,386 | 9,409 | 9,371 |
|  |  | lisboa | 19,496 | 22,658 | 23,331 | 23,864 | 17,870 | 21,301 | 22,475 | 24,337 | 7,526 | 8,587 | 8,548 | 8,573 |
|  |  | norte | 19,457 | 21,515 | 20,996 | 21,884 | 24,165 | 28,376 | 30,211 | 33,108 | 13,436 | 15,034 | 15,048 | 15,065 |

### Prescription and consumption profile by class of psychotropic drugs (2016–2019)

Tables 1–2 and supplementary Figure A1 disclose the consumption rates (DIDs), number of users, and expenditure volumes during the period of analysis for the three classes of psychotropic drugs. Significant growth is observed in the number of users with prescribed antidepressants and antipsychotics (Figure A1a), representing a 20% increase between 2016 and 2019 in both classes – approximately 250,000 new users with prescribed antidepressants and 100,000 new users with prescribed antipsychotics. The number of users with prescribed benzodiazepines is considerably high, 1.5M (>15% of the Portuguese population) and yet does not seem to be in an increasing trend. The progression of the consumption rates (DIDs) per active drug is provided in Figure 1. Figure A2 in appendix complementarily depicts the number of prescribed users per drug. When considering antidepressant prescriptions (Figures 1a and A2a), we find an increasing trend for selective serotonin reuptake inhibitors where sertraline and escitalopram are among the top 3 most prescribed AD as well as alpha-2 antagonists, particularly trazodone and mirtazapine. The prescription of SNRIs, such as venlafaxine and duloxetine also shows an upward trend in contrast to tricyclic antidepressants showing a clear downward trend. Except for amitriptyline, the prescription of dosulepin, maprotiline, trimipramine, nortriptyline, imipramine and pirlindole seems to suggest discontinuation of their use.

**Figure 1.**
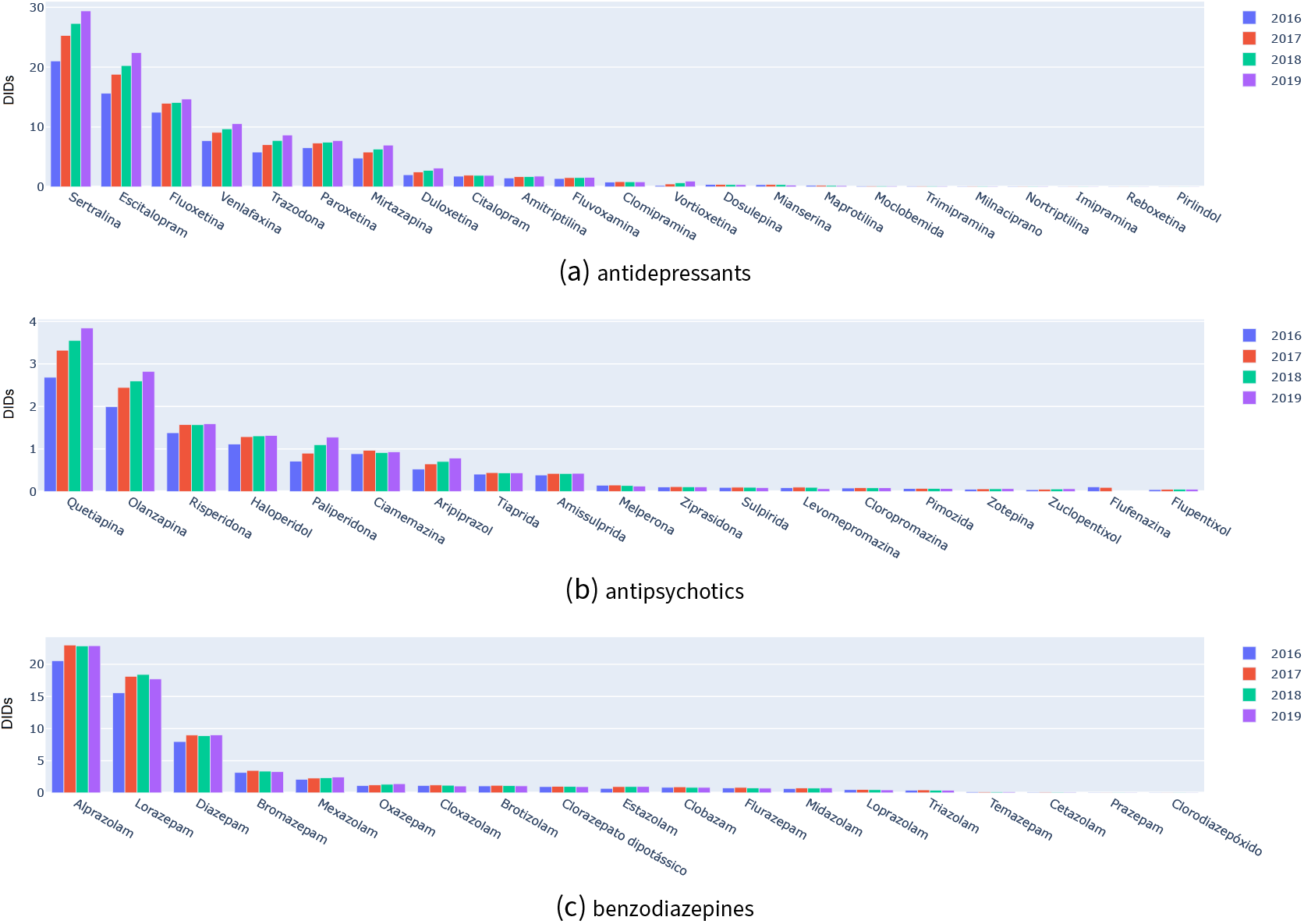
Defined daily dose per 1000 inhabitants-days (DID) per psychotropic drug between 2016 and 2019.

Considering antipsychotics (Figures 1b and A2b), there is a clear prescription tendency towards atypical antipsychotics, with a particular incidence on quetiapine (with an increase of approximately 20,000 users per year), risperidone and olanzapine. Among the typical antipsychotics, amisulpride stands out with a stable prescription rate throughout the study period. The consumption rates of zuclopenthixol, ziprasidone, pimozide, flupentixol and fluphenazine are increasingly residual.

The prescription of benzodiazepines (Figures 1c and A2c) appeared stable in the years we analyzed with a clear preference for the prescription of alprazolam, diazepam, lorazepam and bromazepam, and an increasing trend in the prescription of mexazolam.

### Demographic profiles

The gender and age distribution of consumption rates (Table 1), number of prescribed users (Table 3), and expenditures is visually depicted in the supplementary Figures A3 to A8. We observe a higher incidence of psychotropic drug consumption and prescription in women in all our sub-samples, and significantly higher consumption rates (DID) in the age groups above 40 years (*p*-value<1E-3, quarter estimates). Those at a younger age seem to represent a considerably smaller share of pros and yet appear to be growing expressively (Tables 1 and 3; 18-29 years). Finally, we observed that the volume of expenditure is on par with the incidence of prescriptions in the female population. However, in the male population younger users seem to represent a larger spending per volume of prescriptions. The proportion of antidepressants, antipsychotics and benzodiazepines does not differ significantly by age group. However, there is a predominance of antidepressant use over benzodiazepines in the younger age groups.

Considering the gender distribution per psychotropic drug (Figures A7 and A8), we observe that most drugs do not generally show a significant deviation from gender distribution expectations (*p*-value<1E-3), with few exceptions, including tiapride which is more frequent in men.

### Geographical Distribution

The geographical incidence of consumption rates, prescribed users, and total-and-relative expenditures by group of psychotropic drugs is visually drawn in supplementary Figures A9 and A10. The allocation of users across geographies was inferred from their primary health care centre and not from the place of prescription (as it is often carried out in large centres). The analysis of the geographical distribution of consumption rates (Table 1, Figure A9), standardized by the age distribution of citizens per region, reveals some divergences - the Algarve being the region with the lowest consumption of antidepressants and anxiolytics, and the North of Portugal having the lowest consumption of antipsychotics but the highest consumption of anxiolytics. Regions with incidence of prescription per resident above the average include Coimbra, Évora and Portalegre while Faro and Setubal have incidences below the average. The analysis of the variability observed between quarters confirms the statistical significance of the differences reported (*t* -test, *α* = 1E-3, quarter estimates).

Additionally, the analysis of total expenses and co-payments per district (Figure A10) shows considerable agreement with the volume of users (Figure A9). The Centre region stands out as the one that produces the highest expenditure per user, both in total volume and in the volume of state co-payments. In Porto and Guarda regions, despite the number of users being in line with the average, the volume of reimbursements per resident is below average, due to a lower average number of prescriptions per user.

Figure A11 breaks down the expenditure analysis by district to acquire a finer spatial granularity with the aim of identifying areas with deviating prescription profiles in the Portuguese territory.

### Medical specialty of the Prescriber

An initial view of the consumption and prescription rates, the number of prescribed users, and total volume of prescriptions by medical specialty and class of psychotropic drugs is presented in Figures 2 and A12. For this analysis, all prescriptions made between 2018 and 2019 were considered. First, we observe that over two-thirds of benzodiazepine prescriptions are carried out in General Family Practice, with the sum of all prescribed users by the remaining medical specialties responsible for one third, including psychiatry (11% of prescriptions) and internal medicine (5% of prescriptions). Secondly, and analogously, there is similar representativeness in the prescription of antidepressants, with General Family Practice comprising more than 60% of prescriptions. In this class of psychotropic drugs, psychiatry and neurology specialties have a higher share of prescriptions, representing approximately 20% and 6% of the total volume of prescriptions. Third, General Family Practice is the specialty that most prescribed antipsychotics (47% of prescriptions), followed by psychiatry (34%), which together are responsible for prescribing approximately 80% of antipsychotic prescriptions.

**Figure 2.**
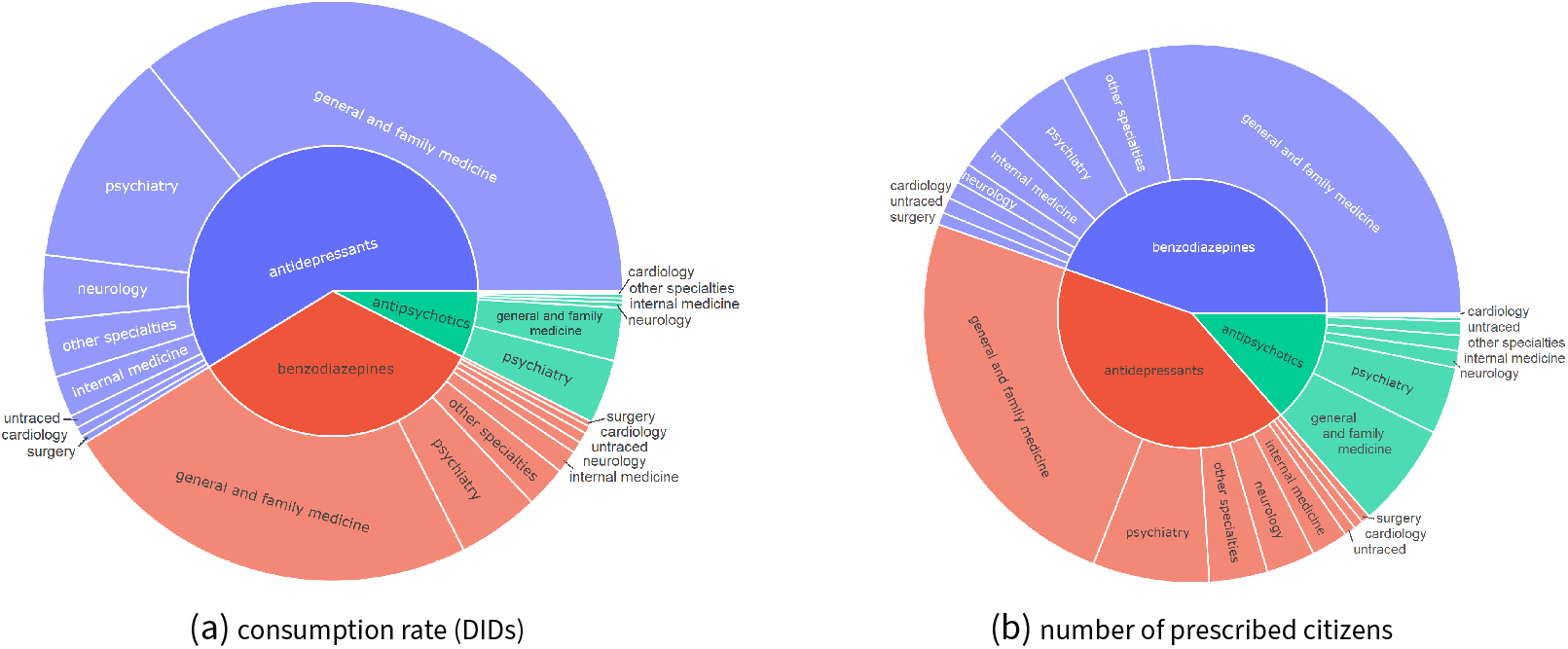
Prescribing medical specialty, 2018–2019.

Figure A13 place the focus on the proportion of psychotropic drug classes by medical specialty. Most medical specialties prescribe more benzodiazepines and a reduced proportion of antipsychotics (less than 10% of total prescriptions in more than 80% of the specialties). Alongside this observation, there are significant variations in the proportion of prescriptions for each class of psychotropic drugs between medical specialties.

Looking in detail at the drugs by group in Figure 3, we can observe that the proportion of the different antidepressants does not differ significantly from specialty to specialty. Nevertheless, we observe that surgery, neurology, orthopaedics and rheumatology prescribe significantly more amitriptyline and duloxetine than the remaining specialties. In turn, psychiatry prescribes some active substances with low prescription in the other specialties, including citalopram, clomipramine, fluvoxamine and vortioxetine.

**Figure 3.**
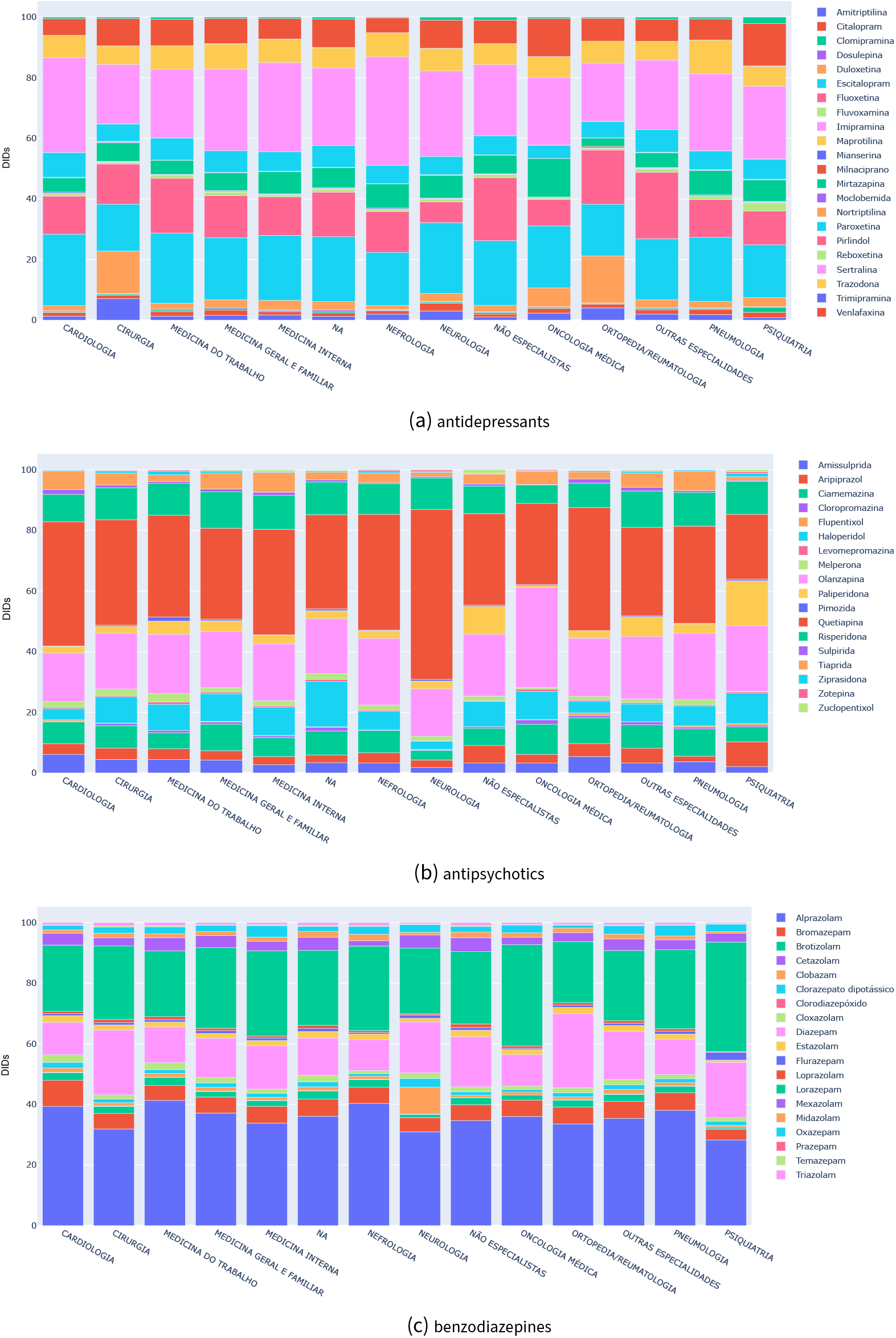
Distribution of psychotropic drugs (DIDs) prescribed per medical specialty, 2018–2019.

In the context of antipsychotic prescriptions, there is also a balanced drug distribution per specialty, with a clear cross-sectional preference for quetiapine. Medical oncology seems to make more relative use of haloperidol than other specialties, which seems to be motivated by its role in reducing nausea and vomiting as a complement to its antipsychotic functions. Similarly to antidepressant prescription, we observed that psychiatry prescribes moderately some antipsychotics, such as aripiprazole and paliperidone, with residual representation in the other specialties.

Finally, in terms of benzodiazepines, we also observed a non-deviating distribution of drugs by specialty. specialties with low prescriptions of benzodiazepines tend to prefer diazepam, while specialties with more active prescription profiles tend to prefer alprazolam. In psychiatry and oncology, the consistent preference for lorazepam also stands out.

## 4 Discussion

Our data suggest an overall increase in the prescription rates, volume of expenditure, number of prescriptions, and cases prescribed over the years — the Portuguese trend is similar to worlwide trends (OECD 2020; Bachhuber *et al*. 2016; Murphy *et al*. 2016; Xu *et al*. 2021; Hálfdánarson *et al*. 2017). This increase was greater between 2016 and 2017, probably reflecting a change in policy regarding the mandatory use of PEM, thus increasing the prescription registry. Among the three drug classes, the antidepressants were the ones that suffered the largest increase during this period, with more 45,13 DID than the ones prescribed in 2016, corresponding to a 47% increase. The last studies on the matter in Portugal reported a 20% increase per year in the antidepressant consumption rates between 2000 and 2012 (Caldas de Almeida *et al*. 2013). This new data shows that this trend seems to be slowing down, yet solidly positive. It is also important to notice that, especially within this class, this trend may have accelerated after 2019 considering the impacts of the COVID-19 pandemic.

When considering the OCED report on antidepressant consumption, our results are in line with the presented estimates up to 2017 (OECD and EU 2018), confirming that Portugal is significantly above the OECD average for antidepressant use (63.3 DID). The continuous increasing trend may result from an improved recognition of mental disorders and accessibility to treatment, including better clinical guidelines. However, it is also likely to be associated with increased societal stressors; the worsening of medical intervention, namely shorter consultation length and lesser frequency; as well as the loss of psychosocial interventions (Tobin, Bury, and Cullen 2020; Johnson *et al*. 2017). SSRIs stand out as the most prescribed antidepressants which are along with the guidelines for depressive and anxiety disorders and evidence from other countries (Bogowicz *et al*. 2021; Dai Cao *et al*. 2021; Livingston 2020). This type of antidepressant, particularly sertraline, is probably preferred by physicians due to its well-documented great combination of efficacy and acceptability (Cipriani 2020). An atypical antidepressant, trazodone, is the fifth most prescribed drug in the country, a fact probably justified by its use as an add-on during adjustment reactions and depressive episodes in users with insomnia. In fact, data from the United States of America show that most trazodone prescriptions are explained by its off-label use for the treatment of primary or secondary insomnia (Stahl 2009). In another stance, tricyclic antidepressants (TCA) seem to have become discontinued, perhaps due to their low safety and tolerability. However, as observed in similar studies (Lockhart and Guthrie 2011; Saragoussi *et al*. 2012), amitriptyline stands out in this scenario, probably, again, derived from its non-FDA approved uses for symptoms and disorders such as insomnia, chronic pain or bladder pain syndrome (Sultana *et al*. 2014; Verhaak, Beurs, and Spreeuwenberg 2019).

When looking at the use of antipsychotics, Portugal also registers a clear upward trend of its prescription, with a 41% increase in this period, especially among atypical antipsychotics, with quetiapine standing out as the most prescribed antipsychotic in Portugal. This can be partly explained by its off-label use for non-approved FDA indications such as anger management, dementia and insomnia (Office of Public Affairs 2010). Despite the risk of metabolic syndrome associated with their use, risperidone and olanzapine are still prescribed to a greater extent than other antipsychotics, possibly due to their key sedative properties and approval in non-psychotic mood disorders. Amisulpride prescription also stands out perhaps due to its use in depressive symptoms in those with assorted mental symptoms (e.g. conversion/somatic), as an add-on for bipolar type-1 disorders (Zangani *et al*. 2021; Carta *et al*. 2006), rather than its sole use as an antipsychotic agent (Nuss, Hummer, and Tessier 2007). As observed with tricyclic antidepressants, several first-generation antipsychotics are no longer part of Portugal prescription choices, probably exposing their side effect profile as D2 antagonists (extrapyramidal effects) or low antipsychotic effect (Buhagiar, Ghafouri, and Dey 2020; Marston *et al*. 2014; Morrens *et al*. 2015; Prah *et al*. 2012; Shah *et al*. 2011). In the transition to second-generation antipsychotics, ziprasidone seems an exception, with a reduction in its prescription, perhaps for alternate drugs choice with better safety and tolerability profile (e.g. vs aripiprazole) and efficacy (e.g. vs olanzapine and risperidone) (Gardner *et al*. 2013). The growing use of antipsychotics could also be explained their increasing use in Bipolar Disorder (Hayes *et al*. 2011), and the growing familiarity of its prescription among GPs, able to manage them better than in the past and are less afraid of using them (Buhagiar, Ghafouri, and Dey 2020; Livingston 2020; Morrens *et al*. 2015).

The prescription of benzodiazepines, desspite the sudden increase in the first year of the study, seem to be stable from 2017 to 2019. This might refer not only to a growing consciousness of potential hazards associated with its use but to the crescent shift to SSRIs (Rosman, Le Vaillant, and Pelletier-Fleury 2011; Šubelj, Vidmar, and Švab 2012). However, it is worth noticing that this overall trend is supported by the decrease in prescriptions among younger adults (18–69 years), while an increase for users above 70 years old is still evident. This is of particular concern as benzodiazepines are considered a potentially inappropriate medication (PIM) in this population and their prescription should be avoided (American Geriatrics Society *et al*. 2015; O’Mahony *et al*. 2018). With respect to the general choice of benzodiazepines, our results go along with previous evidence where alprazolam, diazepam and lorazepam are the most commonly prescribed anxiolytic drugs (Rosman, Le Vaillant, and Pelletier-Fleury 2011; Šubelj, Vidmar, and Švab 2012). Indeed, alprazolam was the most prescribed in our sample, likewise prescription practices of most countries in the world (Ait-Daoud *et al*. 2018). While short half-life benzodiazepines such as alprazolam may represent fewer hazards (Hanlon *et al*. 2002) they are frequently associated with dependence (Cuevas, Sanz, and Fuente 2003). Alprazolam use is seconded by diazepam, a long-acting benzodiazepine with more than 50 years of history, while lorazepam takes up the 3rd place in the prescription profile which is along with its extensive and safe use in situations where there is liver disease/damage (Ghiasi, Bhansali, and Marwaha 2021).

Considering the demographic correlates, although the use of psychotropics at younger groups represent a smaller share, they seem to grow expressively from 2016 to 2019, a trend in line with what’s happening other countries (Jack *et al*. 2020). This can either moved by societal stressors, earlier diagnosis, and/or failure to provide other forms of early intervention (e.g. psychotherapy). This increase is in conformity with previous evidence that considers aggravated societal factors (employment insecurity, low income, reduced social benefits and recession) impacting the mental health in youth (Silva *et al*. 2020). The prescription of psychotropics in subjects over 50 years constitutes more than 50% of the Portuguese total share and, while this can be driven by the ageing of the Portuguese population (Curkovic *et al*. 2016), it can be aggravated by the cumulative effect of the use of psychotropic drugs in adaptive reactions which are kept beyond their actual needs. In all three drug classes, the consumption rates increases with age. With antidepressants and benzodiazepines, we observe a linear progression, with an increase of 49,5 DID of antidepressants and 28,4 DID of benzodiazepines per age group. In the case of antipsychotics use, there is a slow increase from 18 to 49 years which becomes stable until reaching the group above 80 years old, where it suddenly increases by 63,8%, a fact that could be explained by their use in dementia profiles, amongst other conditions (IHME 2022, 2019). While the higher incidence of antipsychotics in the >80 years old group might correspond to actual needs for behaviour control and other neuropsychiatric symptoms (Liperoti, Pedone, and Corsonello 2008), there is evidence alerting to the fact that, in some contexts, only 10% of psychotropic drugs in the elderly are correctly prescribed (Spek *et al*. 2016). Studies and policies to treat mental health disorders in elderly ages are thus fundamental to assess the current status and the role of non-pharmacological intervention on issues such as loneliness that could reduce the use of psychotropic drugs. Considering benzodiazepines, an increase has been observed for young populations in many countries, frequently associated with long-term use patterns against international and national guidelines (Sidorchuk *et al*. 2018). In contrast, in the Portuguese case, a higher proportion of this age group uses antidepressants. While we can hypothesize that the clinicians awareness to paradoxical effects of benzodiazepines in adolescents (Mancuso, Tanzi, and Gabay 2004) or cautions related with the risk of addiction support this observation, we further hypothesize that these results are complementarily driven by the increasing evidence given to the selection of antidepressants as the long term psychotropic choice of treatment for anxiety (Gomez, Barthel, and Hofmann 2018) and depressive disorders (Kendrick, Taylor, and Johnson 2019).

Women represent the group with the highest consumption rates (DIDs) in general, approximately three times more antidepressants, as well as two times more benzodiazepines, than men. The observed gender distribution of psychotropic drug use is supported by previous worldwide (Boyd *et al*. 2015) and Portuguese studies (Silva *et al*. 2020). Discussion on this topic includes the possibility that men could be under-treated and women over-treated with antidepressants (Sundbom *et al*. 2017). While some consider adaptive reactions to be more common in women, the most frequently considered reason for this clear discrepancy is that internalized stigma for help-seeking might hinder the medicalization of suffering in males. Only in the antipsychotic prescriptions do men register higher values with a 14% higher consumption rate. Tiapride use in Alcohol Use Disorders, more prevalent in men (Glantz *et al*. 2020), might explain this fact.

The geographical distribution of prescriptions (adjusted for age) also reveal important discrepancies. The North region records the highest prescription rates of antidepressants and benzodiazepines, but the lowest for antipsychotics. Our results are against recent evidence suggesting heavy use of antipsychotics in rural areas in the north of Portugal (Ramos *et al*. 2021). The Algarve region, on the contrary, always assumes low rates in comparison to other regions. This may reflect a chronic lack of access to primary care in this region, with some users being followed up in central hospitals outside Algarve, as well as an increased dependency of the private sector. As the region with most relative coastal area, the exposure to blue spaces may also play a role in these results. Several previous studies suggest that socioeconomic status (inc. health insurance) can underlie geographical differences in prescription patterns (Dennis *et al*. 2020). Future studies are necessary to outline the determinants for higher prescription in each region and support the implementation of subsequent mitigation strategies.

As seen in Figure 2 on the medical speciality responsible for the prescription, General Practitioners (GPs) preside over all other specialties in all three-drug classes, while psychiatry and neurological specialty are together responsible for only 21% of overall prescriptions. In Portugal, primary care services are a strong component of the National Health Service, with GPs being responsible for the management of non-complex affective disorders, as well as the follow-up of chronic and stable psychiatric disorders. This is along with evidence in other countries where GPs are responsible for most drug prescriptions (Mercier *et al*. 2015; Johnson *et al*. 2017; Svensson, Hedenrud, and Wallerstedt 2019). This observation outlines the need for intervention in the training and clinical support about treatment of mental health and psychotropic drugs among GPs, and the introduction of good practice goals in the annual action plans of the primary care units. Our results for antidepressants show a higher use of citalopram, fluoxetine and sertraline as observed in previous studies (Johnson *et al*. 2017) and there seems to be evidence of off-label prescription (Rubio-Valera *et al*. 2012). GPs seldomly use recent psychotropics (Svensson, Hedenrud, and Wallerstedt 2019) or else rely on direct marketing strategies to identify new treatments (Mercier *et al*. 2011) which are adopted under the premise that these new drugs are more effective (Svensson, Hedenrud, and Wallerstedt 2019). Although the use of benzodiazepines and antidepressants is not restricted to the treatment of mental disorders, our figures also raise important considerations regarding the monitoring of users prescribed for mental disorders by other medical specialities, including psychiatry, neurology, and surgery. Short and long term treatment with benzodiazepines might risk being outdated or beyond rationale from the National Institute for Health and Care Excellence (NICE) if not validated by proper specialities (e.g. neurology and psychiatry). Liaison psychiatry should play an important role here. We further observed that surgery, neurology, orthopaedics and rheumatology prescribe significantly more amitriptyline and duloxetine than the remaining specialities, an observation that is associated with the role of these drugs in neuropathic pain control and enuresis (Reddy and Patt 1994). The case of antipsychotics use, which FDA and EMEA approved for symptomatic treatment of psychoses and affective disorders, the extensive prescription rate by GPs could constitute either an off-label use for insomnia or behavioural symptoms, as well as the result of a long term treatment of stabilized effective schizophrenia spectrum disorders under the care of a GP.

### Limitations

First, our study excludes non-benzodiazepine anxiolytic drugs such as zolpidem, limiting our results to the class of benzodiazepines, a care to be undertaken when establishing comparisons against DID references in other countries and historical estimates in Portugal. Although DDD and DID are still considered to be the reference indicators for pharmacoepidemiology studies alike, they can fail to capture the micro tendencies of each prescription and the diagnosis it serves. Due to multi-level privacy concerns, several clinical inputs (e.g. diagnosis information) could not be collect at a nationwide level, which would be pivotal to understand the psychiatric dynamics that go along with the depicted trends, thus preventing us to inquiry whether the changes in prescription and consumption are related with the increasing prevalence of mental illness. Despite our care in identifying geographical residence of individuals by primary healthcare service (the selected coarse-grained territorial division already mitigates this bias), several geographic discrepancies may refer to centralized mental healthcare treatment in Portugal. In addition, we were not provided with complete prescription registry from the autonomous regions of Azores and Madeira due to the non-mandatory use of electronic registry in these regions throughout the period of the cohort study. The inclusion of these regions is expected in the future for an all-encompassing analysis of the country. Finally, there is a remnant use of paper prescriptions up to 2017, nevertheless credited by the Health Ministry as being largely residual and thus not determinant for analysis or discussion.

## 5 Conclusions

This study is the first comprehensively examining and discussing the prescription patterns of psychotropic drugs in Portugal across years 2016-2019. Several of the acquired results are in line with the existing body of research in other countries – an increase in consumption rates and volume of expenditure, as well as a consolidation trend towards the prescription of specific groups of psychotropic drugs. In particular, we observed an increase in the consumption (>30%) and expenditure (37M Eur) of antidepressants and antipsychotics between 2016 and 2019, and a stabilization on the use of benzodiazepines with an overall consumption by >15% of the Portuguese population. With few exceptions, tricyclic antidepressants and typical antipsychotics seem to be under discontinuation suggesting prescribers are favouring SSRIs and atypical drugs. The use of psychotropic drugs seems to be higher in those of older age and in women, and disparate among different Portuguese regions (after correction for age). Sociodemographic, geographical and cost correlates are explored, unraveling relevant drivers to assess the status of psychotropic drug prescription. The analysis of the responsible medical specialities reveals that General Medicine is responsible for approximately 64% of prescriptions, and is arguably less assorted in the choices when considering the changing in the preferred psychotropic drugs by psychiatry and neurology.

Unique aspects to the Portuguese case are also noted and discussed, including the ageing of the Portuguese population, the follow up of mental disorders in GP, and the economic recession. Our findings ultimately constitute opportunities for public health initiatives, the design of new practice recommendations for psychotropic drug prescription, medical training programs, the strenghtening of protocols between psychiatrists and GPs, as well as psychosocial interventions.

We aim to further assess the patterns of psychotropic drug prescription in Portugal by continuing with the collection and processing of PEM data for the subsequent years, estimating the potential impact yield throughout and after the COVID 19 pandemic in the consumption status. In addition, we expect to source complementary clinical data (e.g., diagnosis and interventions) that can be crisscrossed with the available data to understand the underlying rationale for the different patterns of prescription, and set tailor made protocols for deprescribing strategies. As our study further suggests that several prescription patterns, either for their possible long-term or off-label use, should be closely monitored, we further aim to analyze the prescription registries to study mal-prescription patterns, including problems of poliprescription.

## Data Availability

Access to national prescription data acquired from the Electronic Prescription Platform (PEM) is possible under explicit authorization by the Shared Services Ministry of Health (SPMS), https://www.spms.min-saude.pt/contactos.

## Acknowledgments

The authors thank Serviços Partilhados do Ministério da Saúde (SPMS) for the valuable data provision and support. This research work was further supported by national funds through Fundação para a Ciência e Tecnologia (FCT) under INESC-ID pluriannual (UIDB/50021/2020).

### Appendix

#### A1 Complementary results

**Figure A1.**
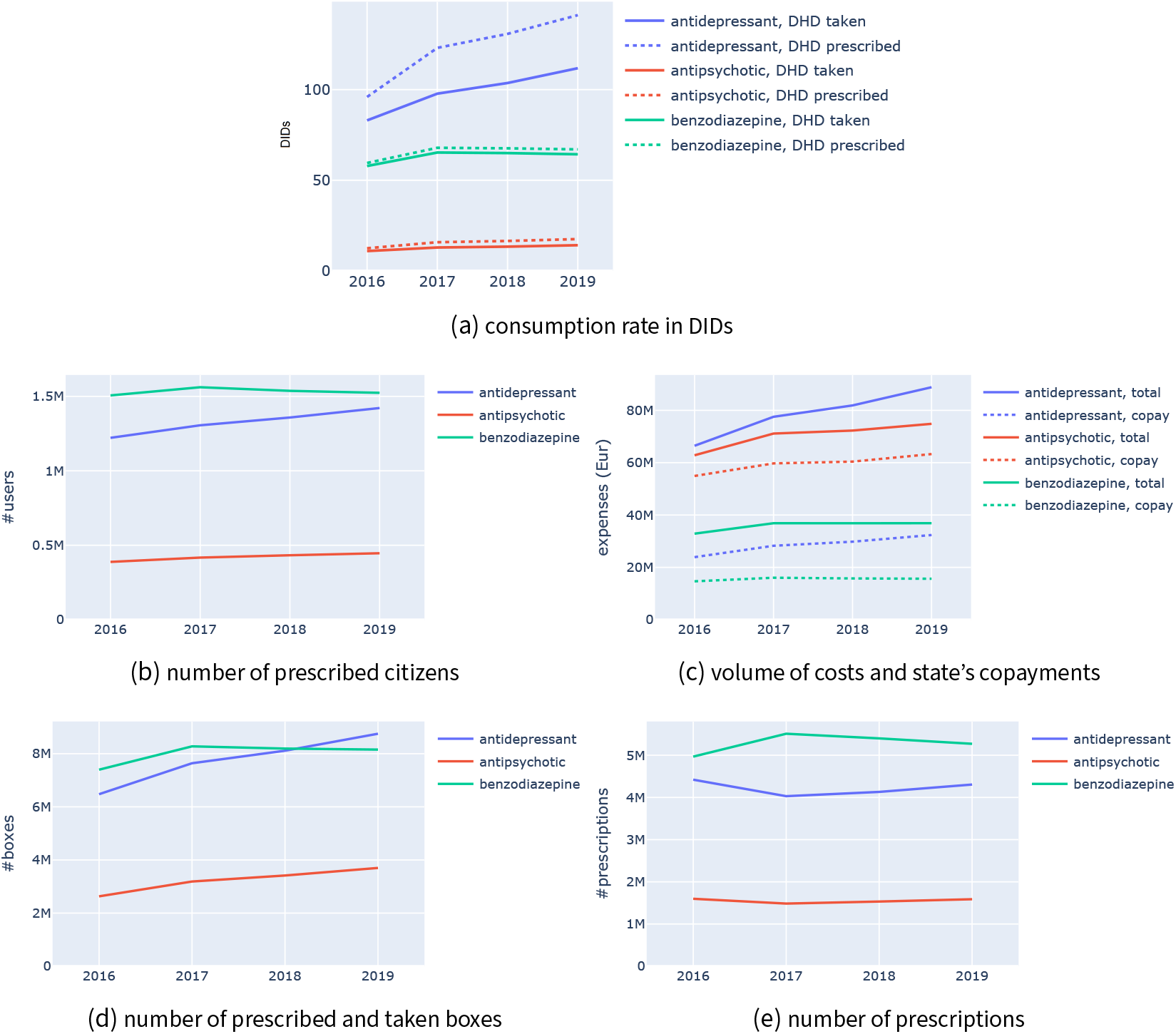
Yearly progression of prescription indicators per psychopharmaceutical drug class between 2016 and 2019.

**Table A1.** Summary of the psychotropic drug prescription status in Portugal: prescription rates across demographic variables expressed in Defined Daily Dose per 1000 inhabitants-days (DIDs).

|  |  | antipsychotics |  |  |  | antidepressants |  |  |  | benzodiazepines |  |  |  |
| --- | --- | --- | --- | --- | --- | --- | --- | --- | --- | --- | --- | --- | --- |
|  |  | 2016 | 2017 | 2018 | 2019 | 2016 | 2017 | 2018 | 2019 | 2016 | 2017 | 2018 | 2019 |
| prescription rates (DIDs) | all | 12.48 | 15.82 | 16.51 | 17.56 | 95.97 | 123.04 | 130.76 | 141.09 | 59.53 | 67.91 | 67.62 | 67.04 |
|  | gender |  |  |  |  |  |  |  |  |  |  |  |  |
|  | F | 11.72 | 15.00 | 15.75 | 16.70 | 140.53 | 179.20 | 189.68 | 203.91 | 79.55 | 90.54 | 89.77 | 88.72 |
|  | M | 13.31 | 16.72 | 17.35 | 18.51 | 46.77 | 61.04 | 65.72 | 71.74 | 37.44 | 42.94 | 43.18 | 43.10 |
|  | age |  |  |  |  |  |  |  |  |  |  |  |  |
|  | [18-29] | 5.83 | 7.41 | 7.93 | 8.64 | 20.15 | 26.41 | 28.71 | 32.45 | 6.44 | 7.18 | 7.06 | 7.08 |
|  | [30-39] | 10.76 | 12.95 | 12.78 | 13.13 | 49.07 | 59.43 | 59.72 | 61.86 | 21.66 | 23.01 | 21.72 | 20.65 |
|  | [40-49] | 16.78 | 20.76 | 21.17 | 22.19 | 95.37 | 120.43 | 125.75 | 133.44 | 50.77 | 56.73 | 55.60 | 54.47 |
|  | [50-59] | 19.18 | 24.23 | 24.98 | 26.65 | 139.25 | 175.51 | 182.68 | 194.32 | 84.53 | 95.98 | 94.97 | 93.33 |
|  | [60-69] | 17.22 | 22.08 | 23.31 | 25.00 | 178.54 | 226.32 | 240.07 | 257.09 | 117.79 | 134.76 | 134.06 | 132.16 |
|  | [70-79] | 16.45 | 21.23 | 22.84 | 24.32 | 211.30 | 274.11 | 295.33 | 323.00 | 144.39 | 164.93 | 165.40 | 165.34 |
|  | [+80] | 24.72 | 33.47 | 36.72 | 39.84 | 221.07 | 303.36 | 339.29 | 378.73 | 166.80 | 198.09 | 202.74 | 206.02 |
|  | region |  |  |  |  |  |  |  |  |  |  |  |  |
|  | alentejo | 12.71 | 16.65 | 17.93 | 18.89 | 93.38 | 117.53 | 125.81 | 133.91 | 47.02 | 52.64 | 51.38 | 50.60 |
|  | algarve | 11.61 | 14.72 | 15.92 | 17.61 | 59.59 | 80.10 | 86.14 | 94.09 | 36.32 | 42.57 | 43.37 | 44.57 |
|  | centro | 13.85 | 17.10 | 17.75 | 18.77 | 102.25 | 126.78 | 133.28 | 142.47 | 64.46 | 72.81 | 72.61 | 71.41 |
|  | lisboa | 13.32 | 17.52 | 18.37 | 19.35 | 92.29 | 123.25 | 132.01 | 142.69 | 44.15 | 51.35 | 50.87 | 50.72 |
|  | norte | 11.10 | 13.77 | 14.23 | 15.30 | 99.53 | 126.78 | 134.73 | 146.51 | 73.86 | 84.11 | 84.09 | 83.46 |

**Table A2.** Volume and share of prescriptions per medical specialty and psychotropic drug class (2018–2019).

| speciality | all |  | antipsychotics |  | antidepressants |  | benzodiazepines |  |
| --- | --- | --- | --- | --- | --- | --- | --- | --- |
|  | volume | share | volume | share | volume | share | volume | share |
| cardiology | 231977 | 1.0% | 11848 | 0.4% | 69892 | 0.8% | 149694 | 1.4% |
| general and family medicine | 14089362 | 63.2% | 1479899 | 47.4% | 5083039 | 60.2% | 7483668 | 70.1% |
| internal medicine | 998272 | 4.5% | 168474 | 5.4% | 350966 | 4.2% | 474543 | 4.5% |
| medical oncology | 90876 | 0.4% | 7472 | 0.2% | 31451 | 0.4% | 51744 | 0.5% |
| nephrology | 63683 | 0.3% | 5520 | 0.2% | 20006 | 0.2% | 38085 | 0.4% |
| neurology | 832563 | 3.7% | 160326 | 5.1% | 469398 | 5.6% | 202146 | 1.9% |
| non-specialists | 168933 | 0.8% | 25117 | 0.8% | 56172 | 0.7% | 86677 | 0.8% |
| occupational medicine | 114671 | 0.5% | 11885 | 0.4% | 40304 | 0.5% | 62053 | 0.6% |
| orthopedics/rheumatology | 151611 | 0.7% | 6610 | 0.2% | 59932 | 0.7% | 84736 | 0.8% |
| pneumology | 95056 | 0.4% | 9254 | 0.3% | 32411 | 0.4% | 53154 | 0.5% |
| psychiatry | 3966556 | 17.8% | 1055735 | 33.8% | 1719096 | 20.4% | 1188595 | 11.1% |
| surgery | 211929 | 1.0% | 20261 | 0.6% | 81027 | 1.0% | 109120 | 1.0% |
| other specialties | 856131 | 3.8% | 86177 | 2.8% | 282776 | 3.4% | 471762 | 4.4% |
| non specified | 430540 | 1.9% | 71625 | 2.3% | 140771 | 1.7% | 215403 | 2.0% |

**Figure A2.**
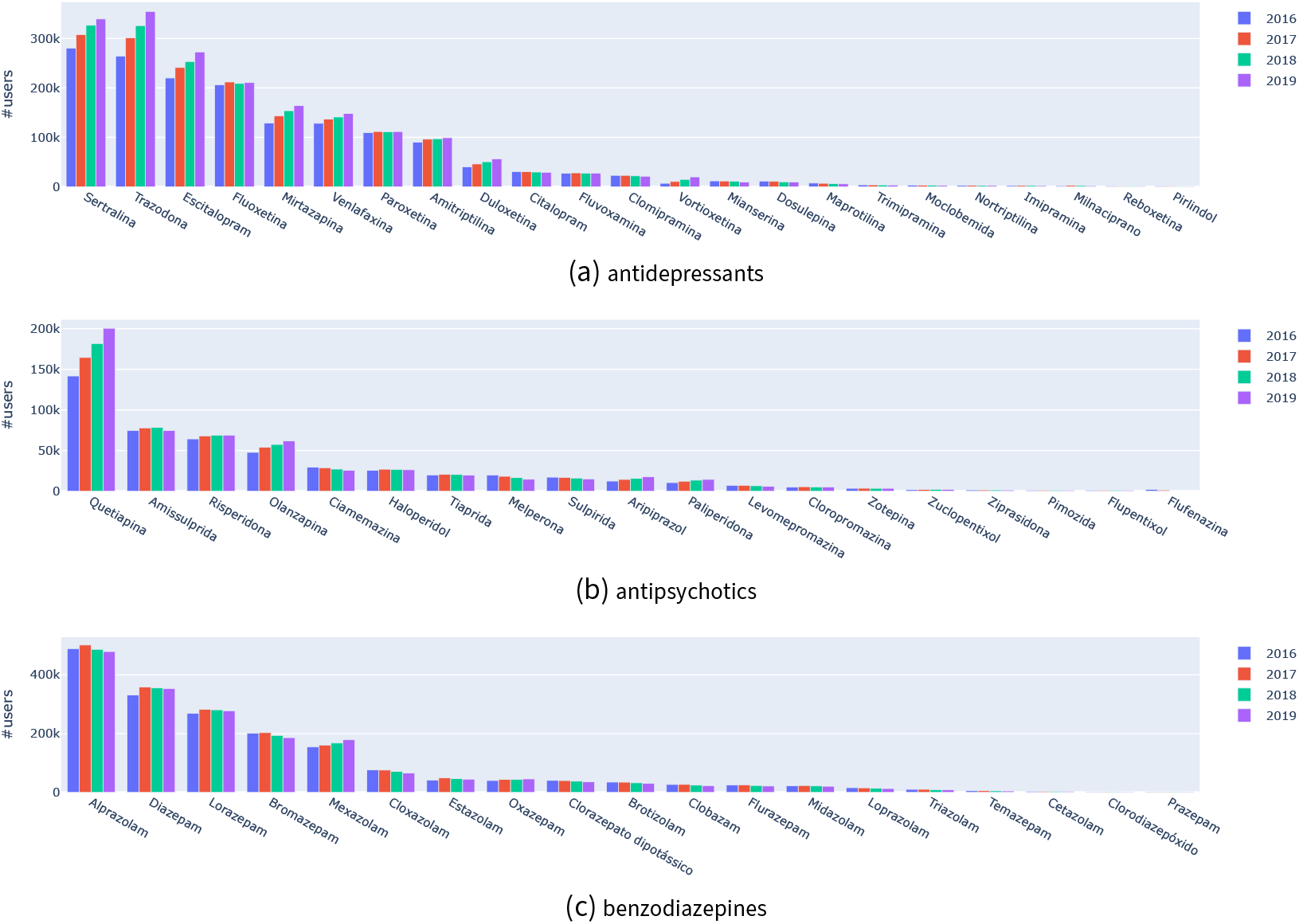
Number of prescribed citizens per psychotropic drug between 2016 and 2019.

**Figure A3.**
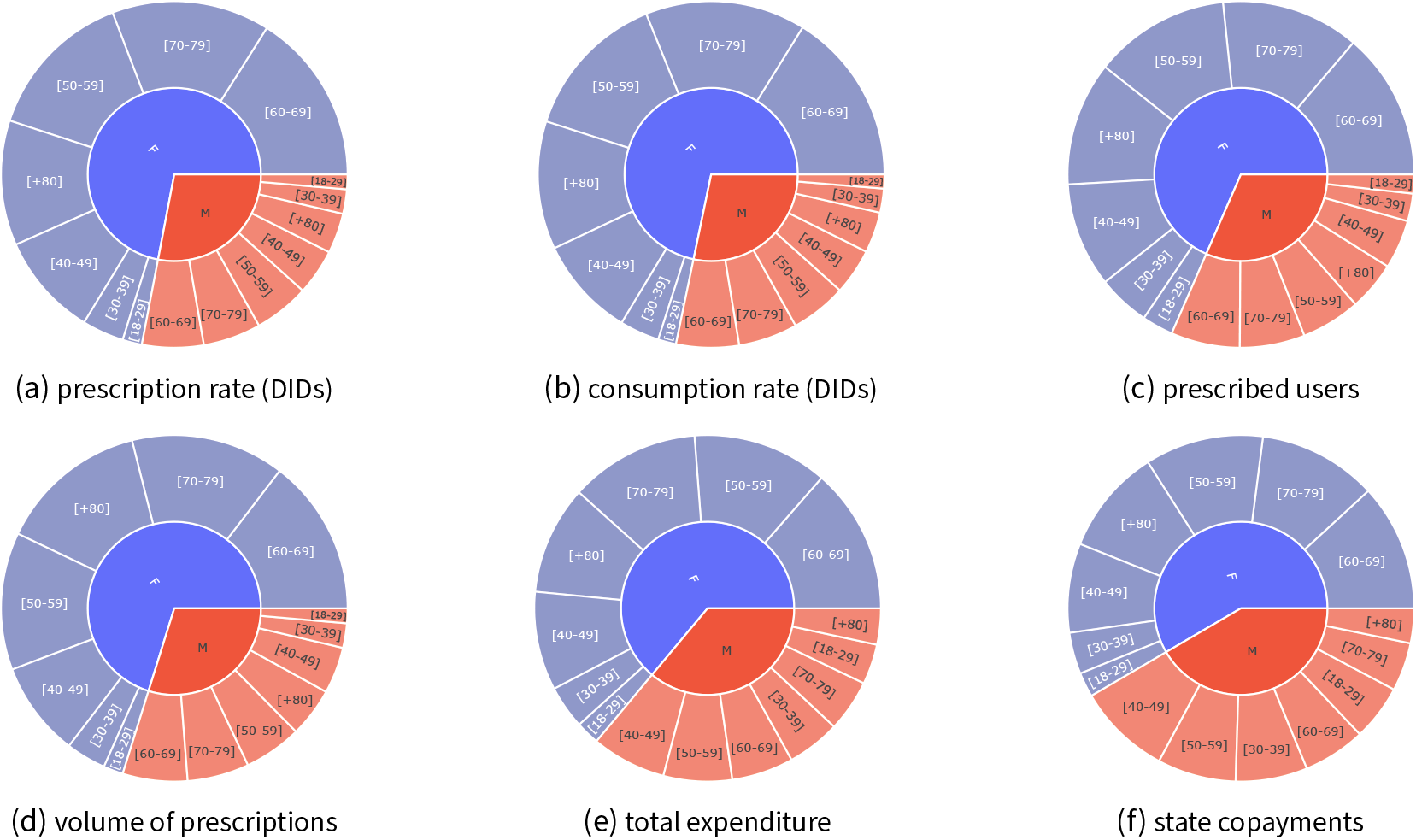
Distribution of psychotropic drug prescription volume across gender and age.

**Figure A4.**
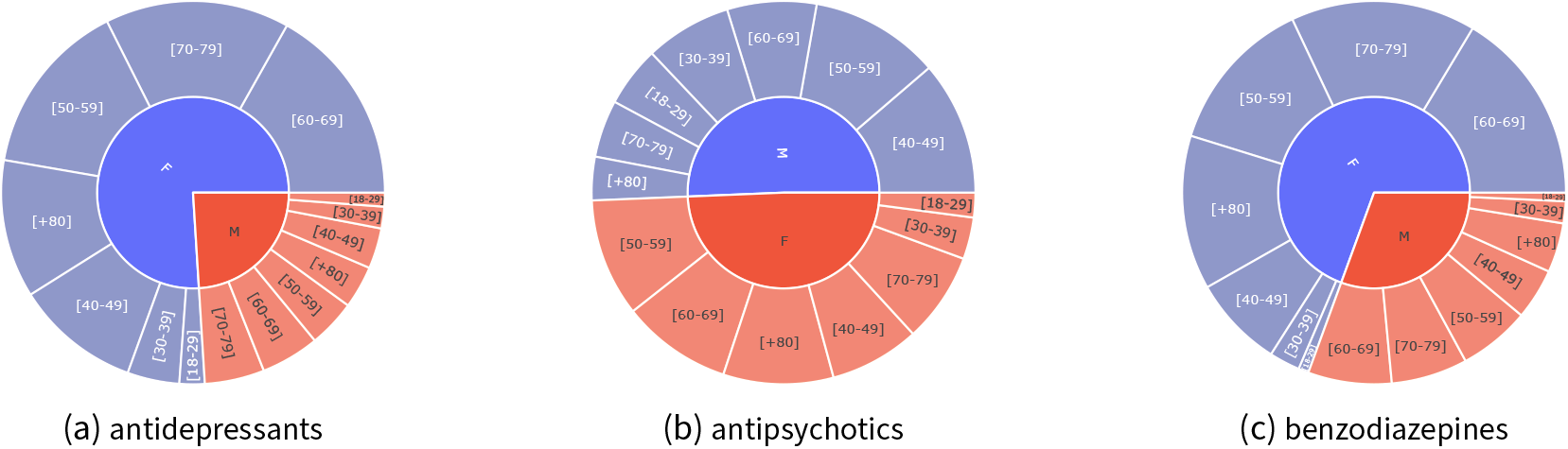
Distribution of consumption rates (DIDs) per drug class across gender and age.

**Figure A5.**
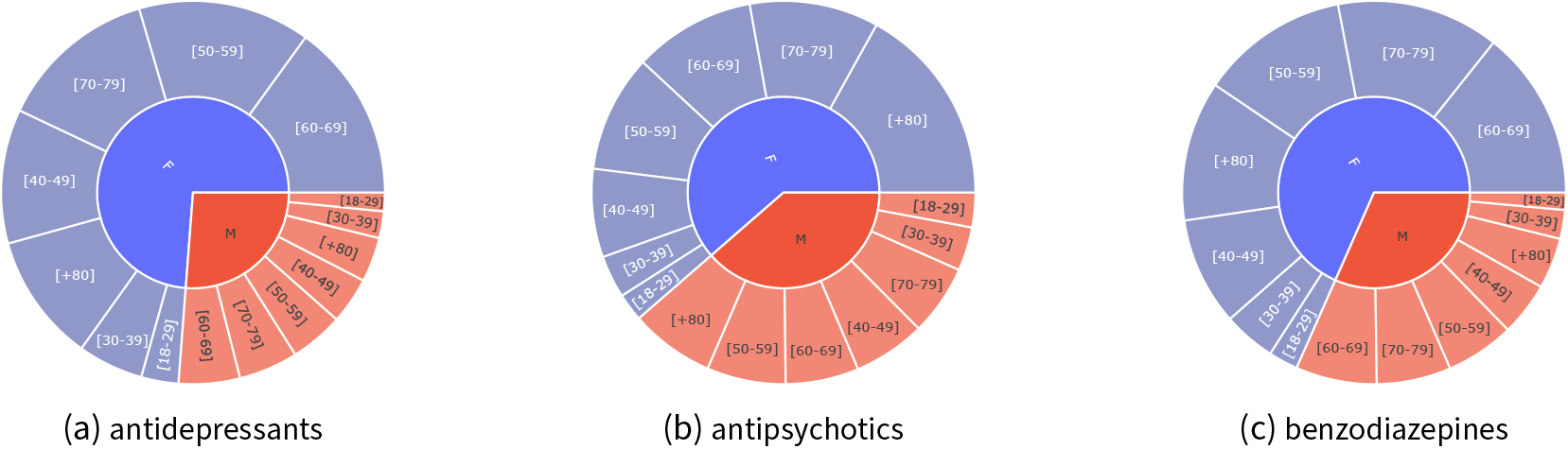
Distribution of the number of prescribed citizens per drug class across gender and age.

**Figure A6.**
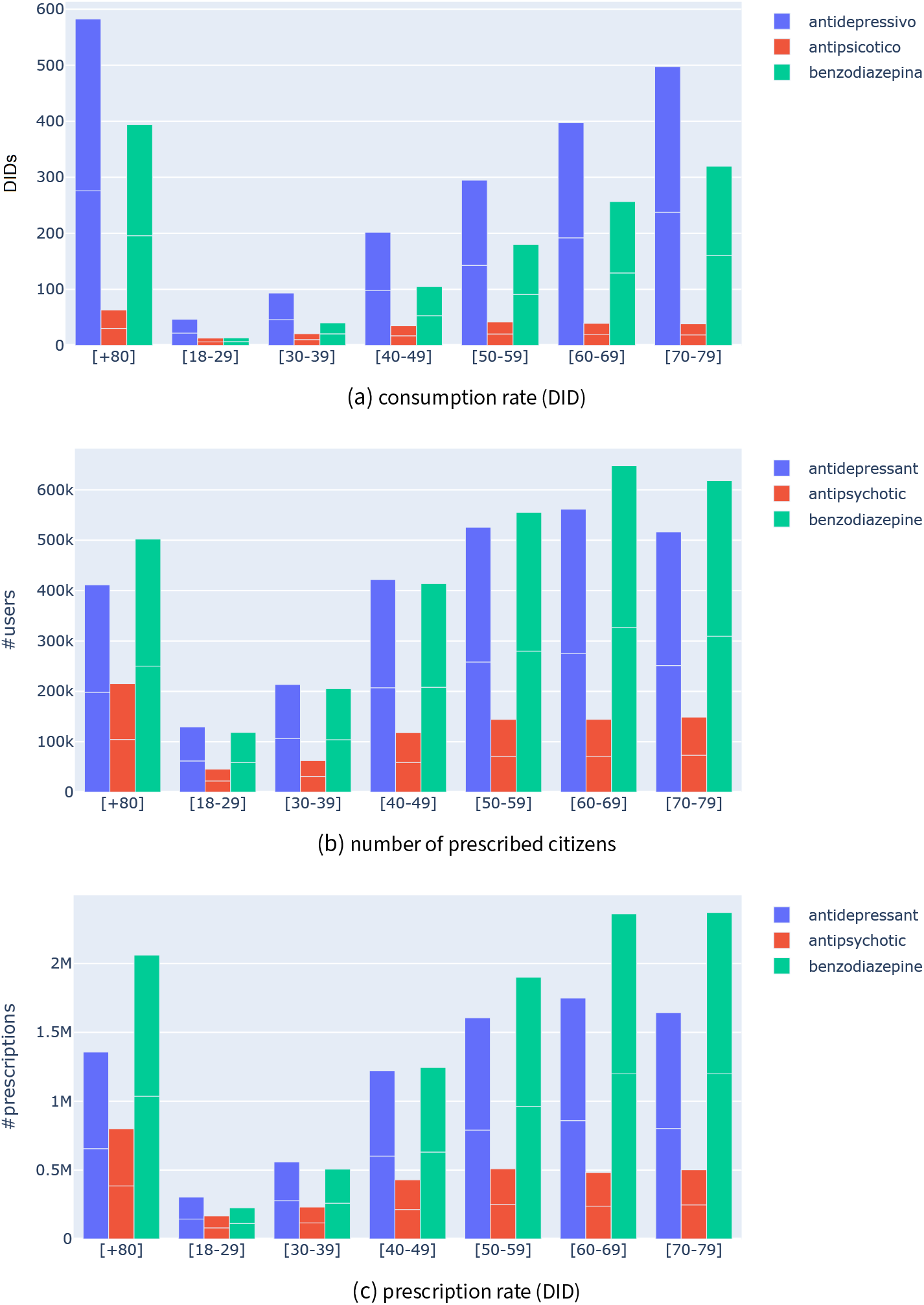
Distribution of psychotropic drugs per age, 2018–2019

**Figure A7.**
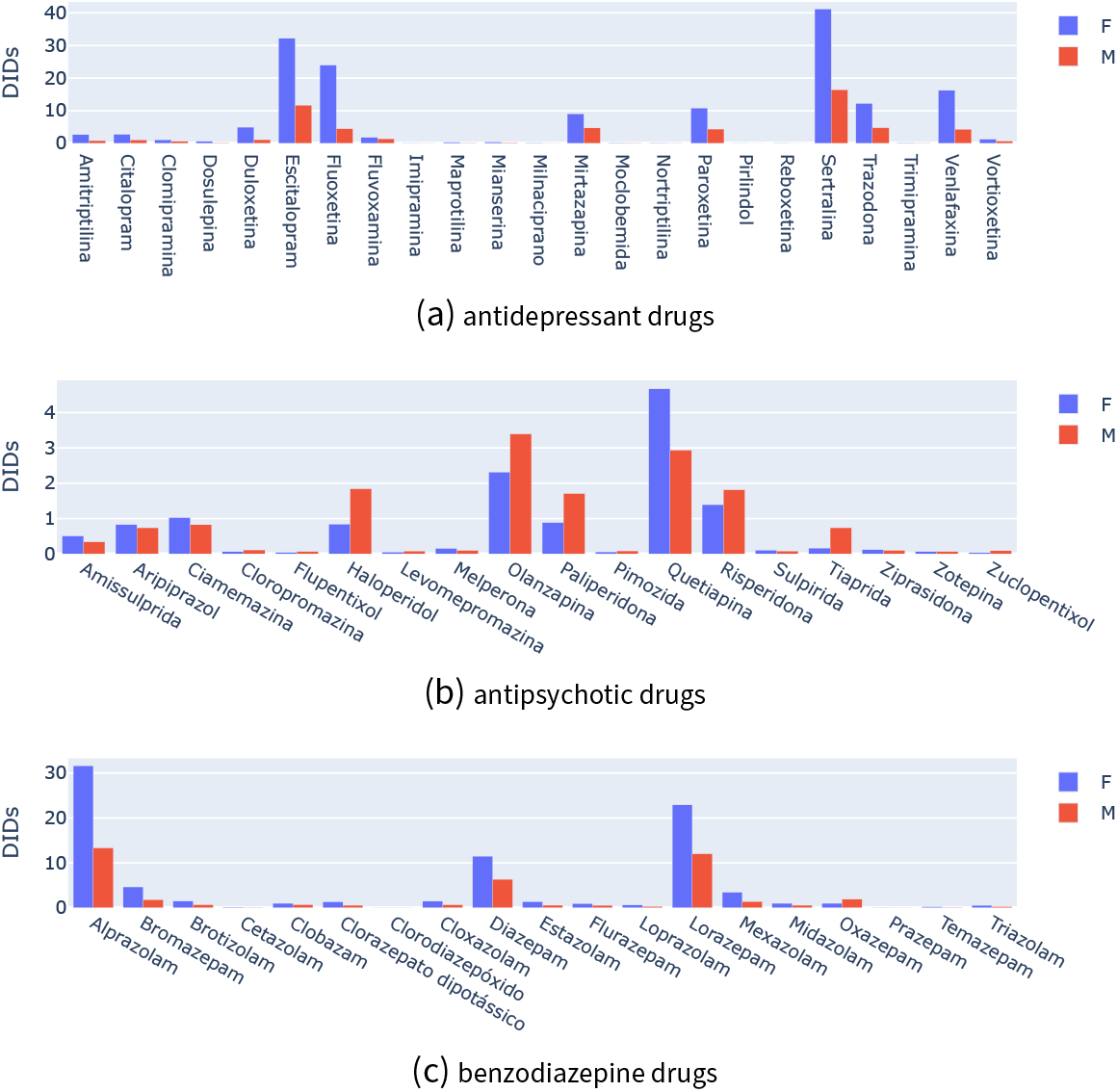
Consumption rate in DIDs per drug and gender, 2018–2019.

**Figure A8.**
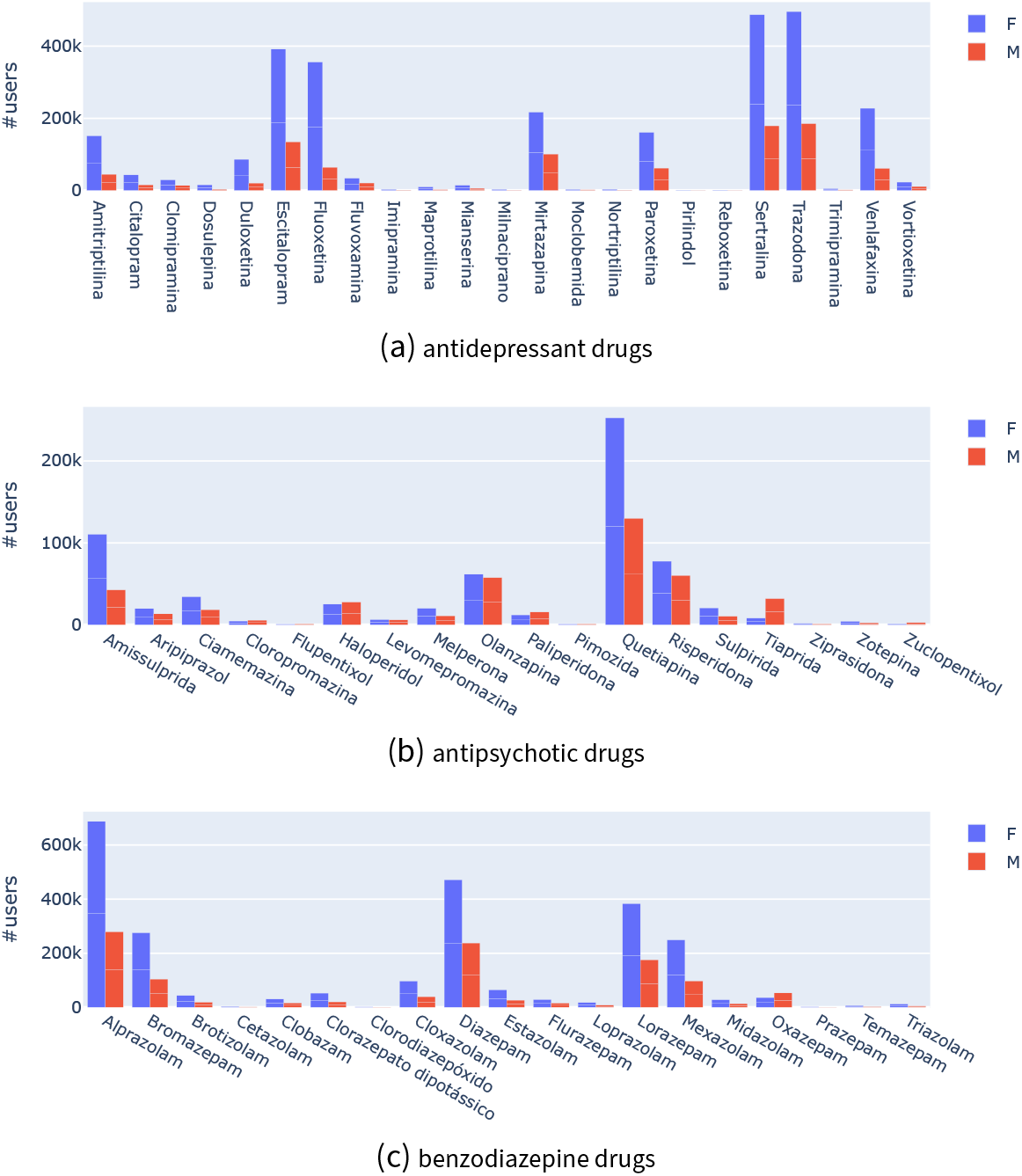
Number of prescribed individuals per drug gender, 2018–2019.

**Figure A9.**
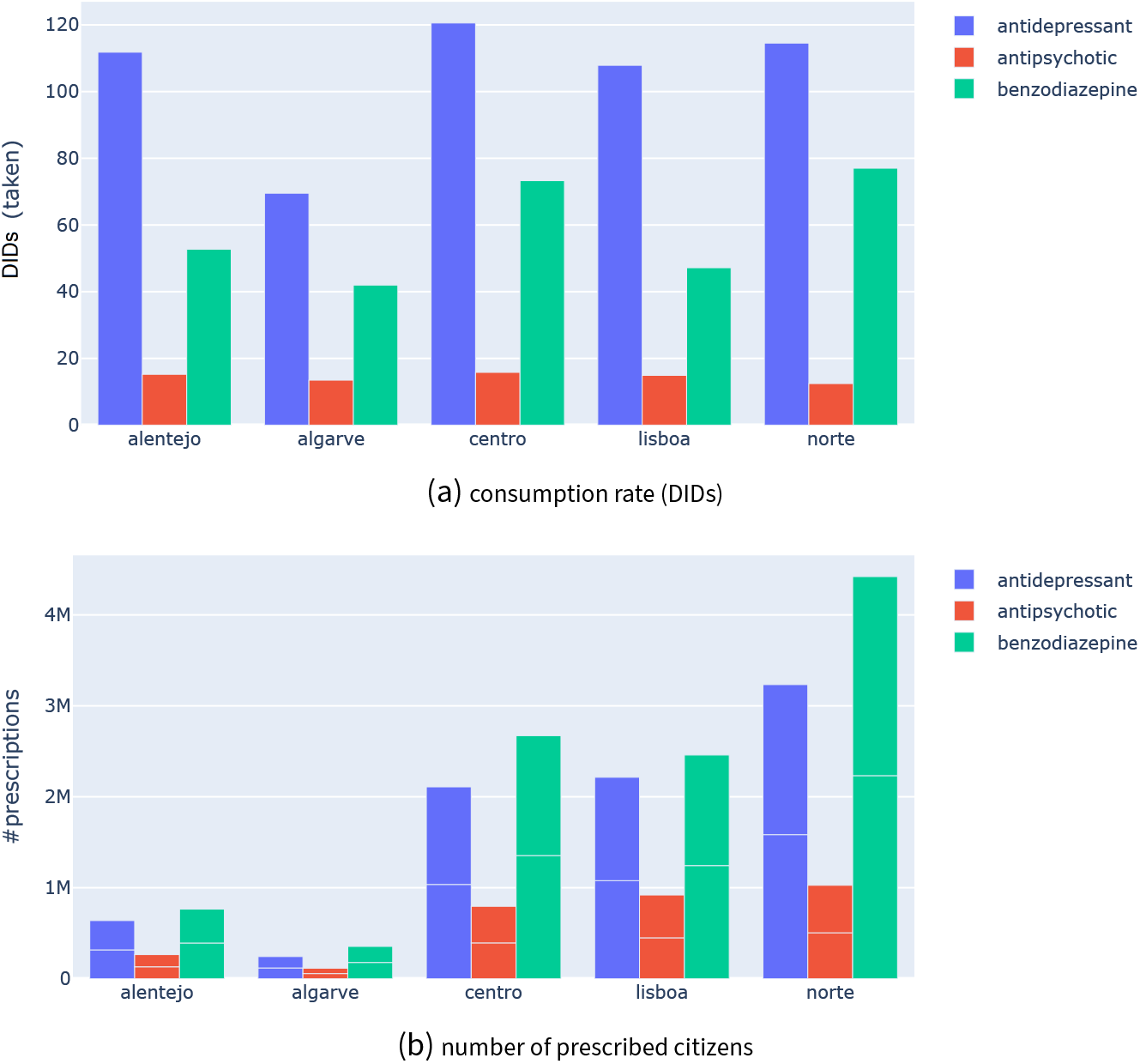
Drug prescription profile per geography after standardization for age groups against census data, 2018–2019.

**Figure A10.**
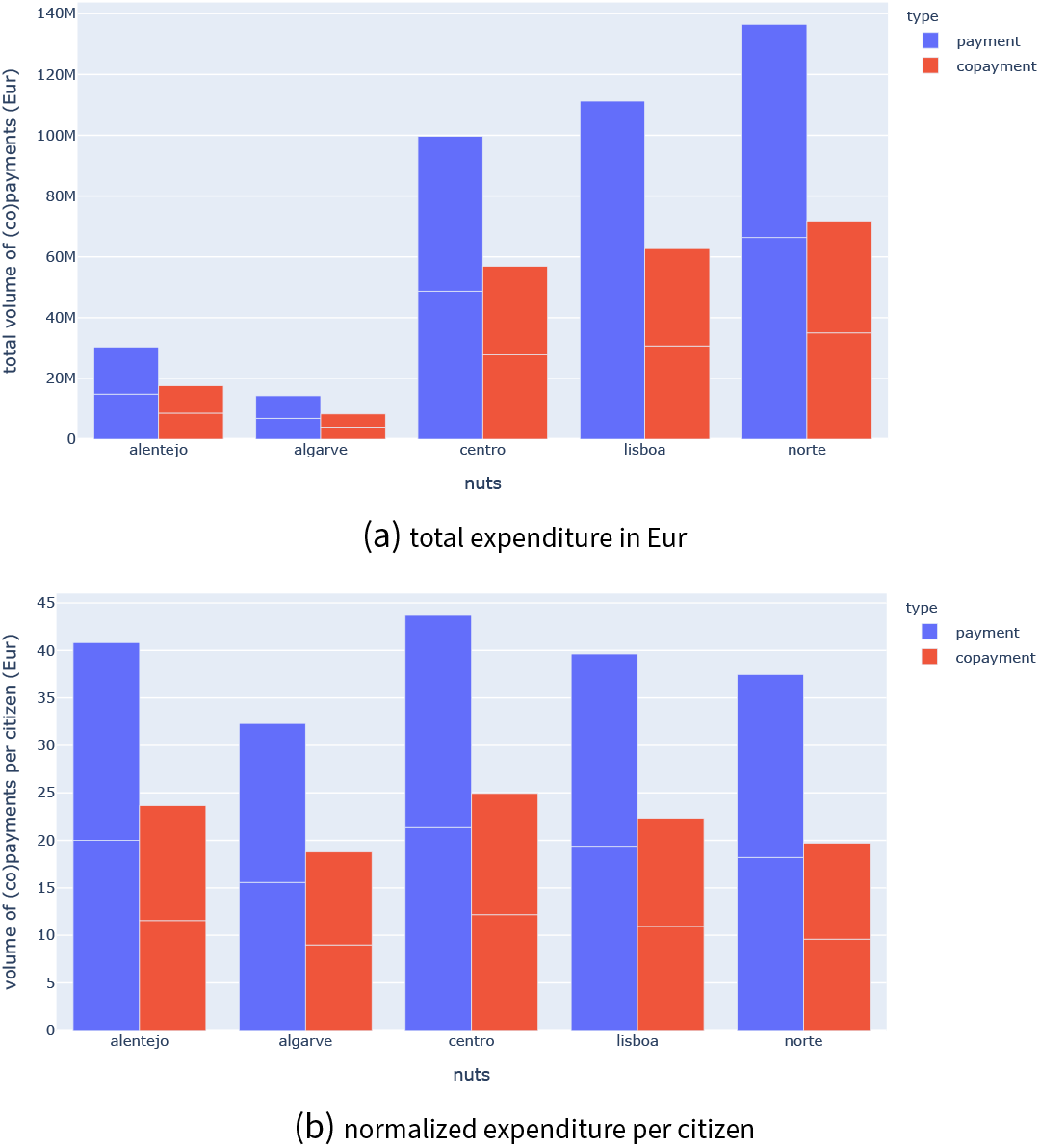
Annual expenditure per geography, 2018–2019.

**Figure A11.**
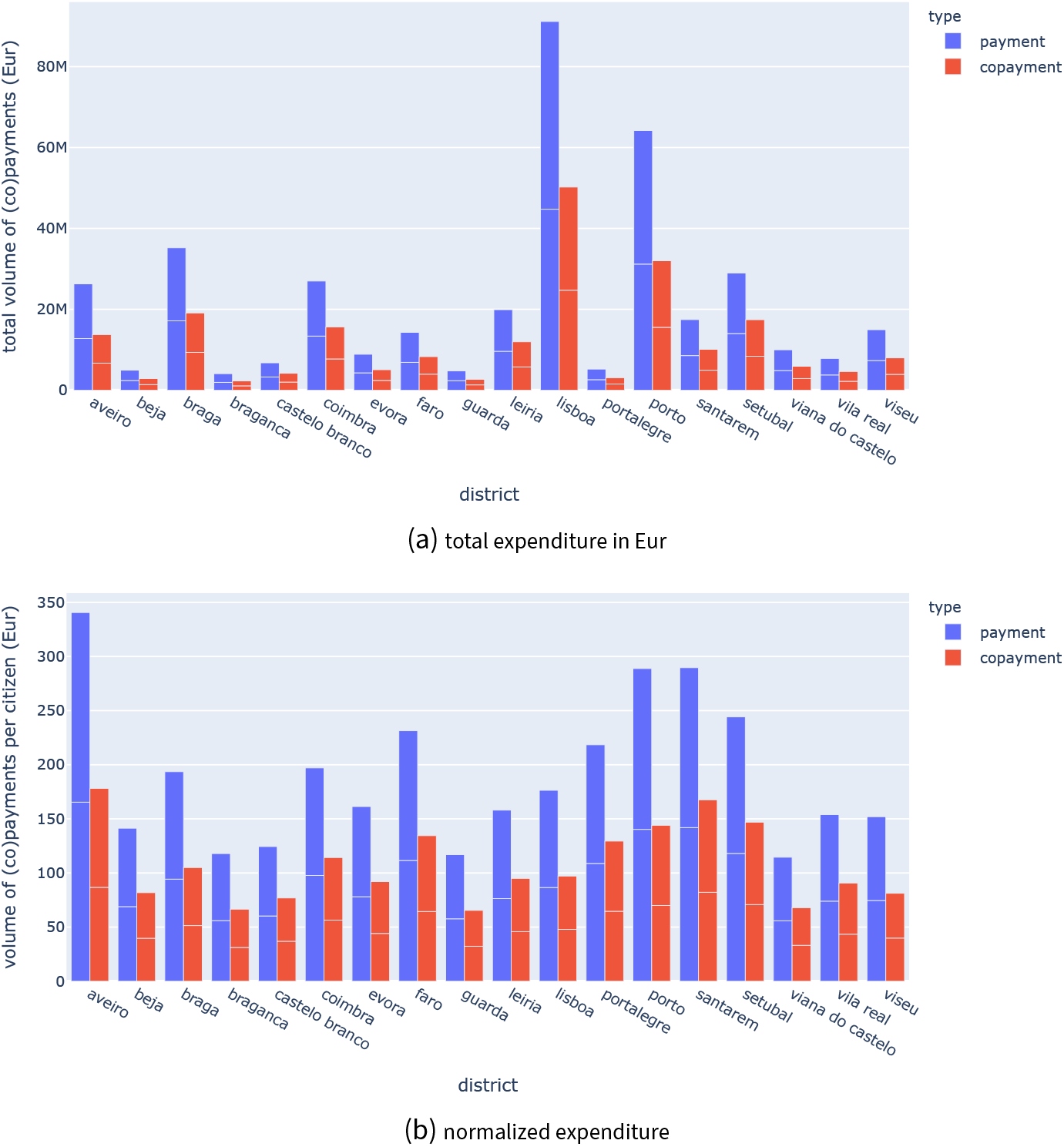
Annual expenditure per district, 2018–2019.

**Figure A12.**
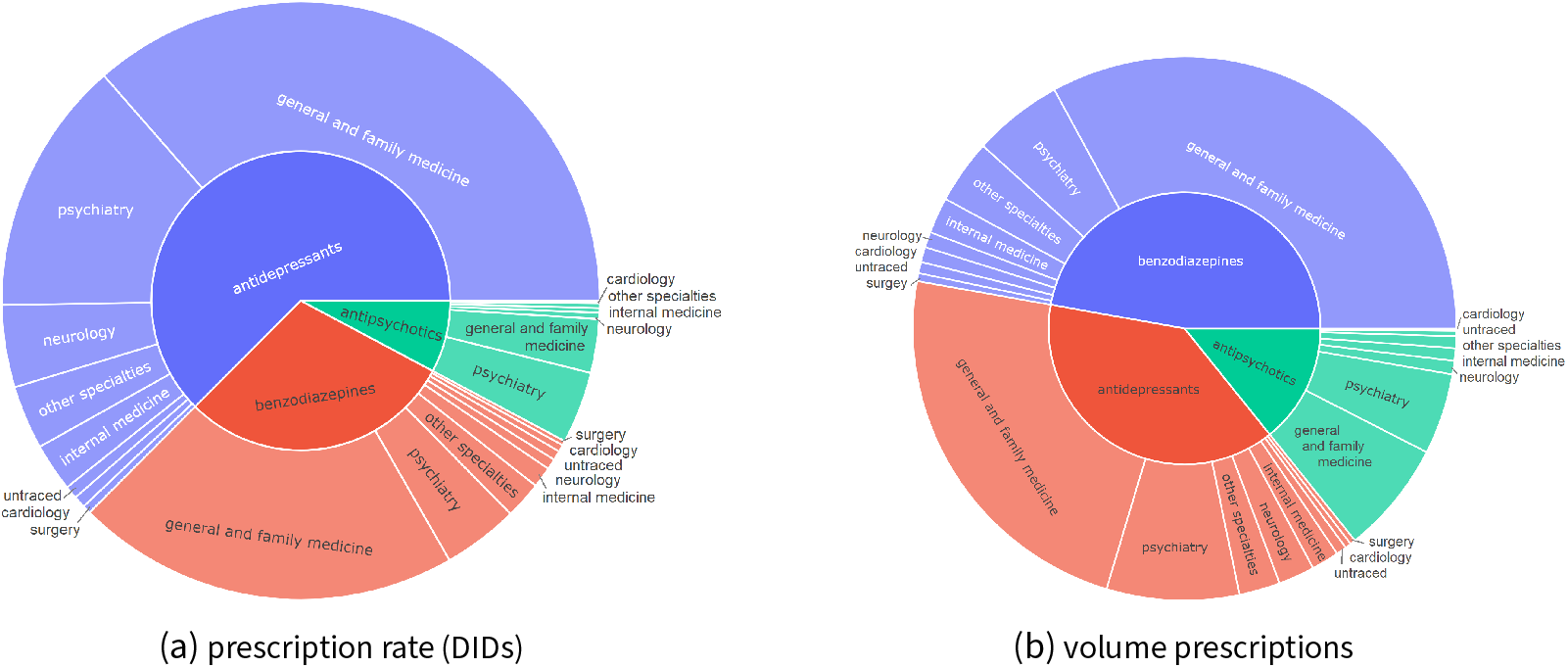
Prescribing medical specialty, 2018–2019.

**Figure A13.**
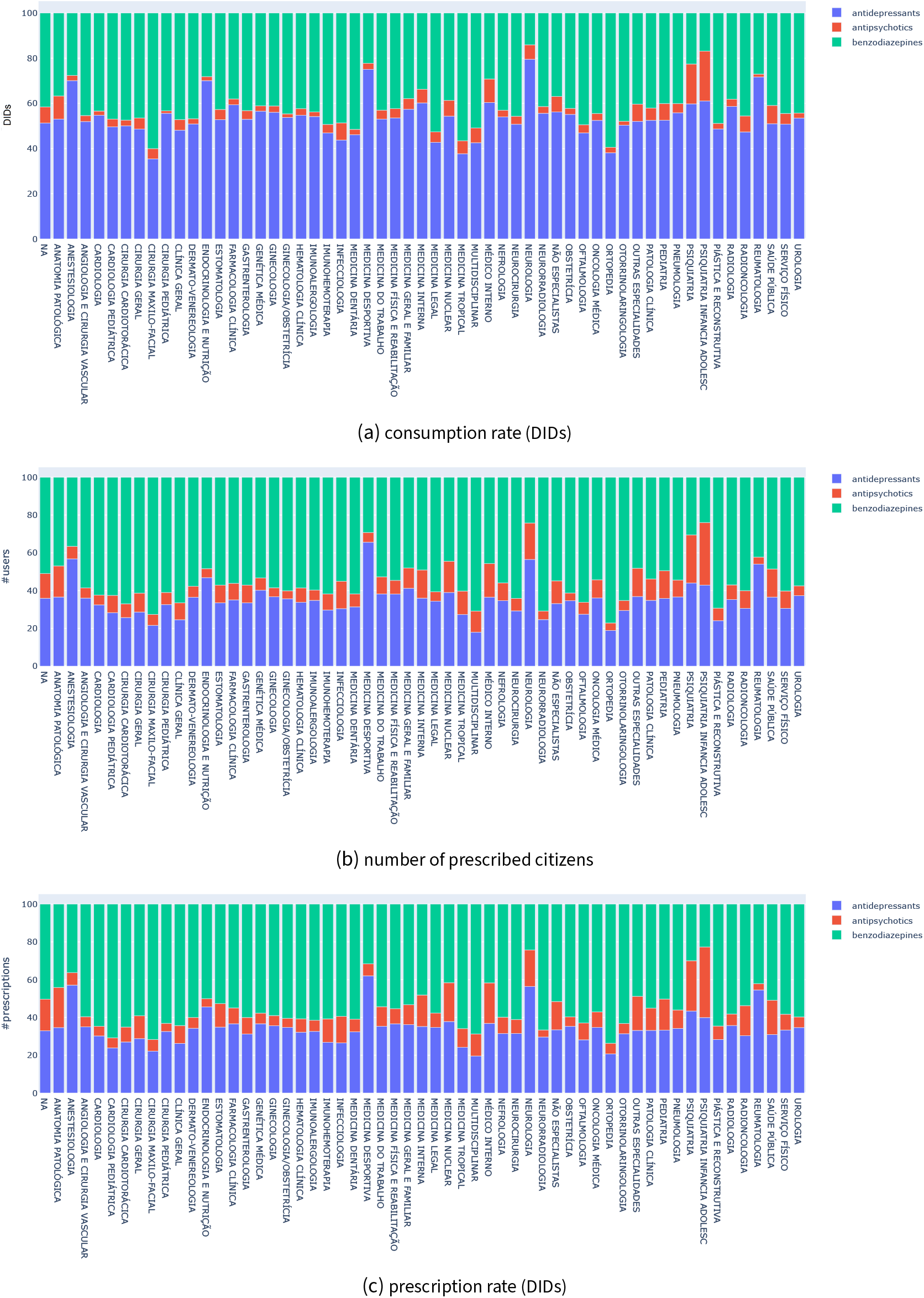
Distribution of the prescription volume for each class of psychotropic drugs per medical specialty, 2018–2019.

**Figure A14.**
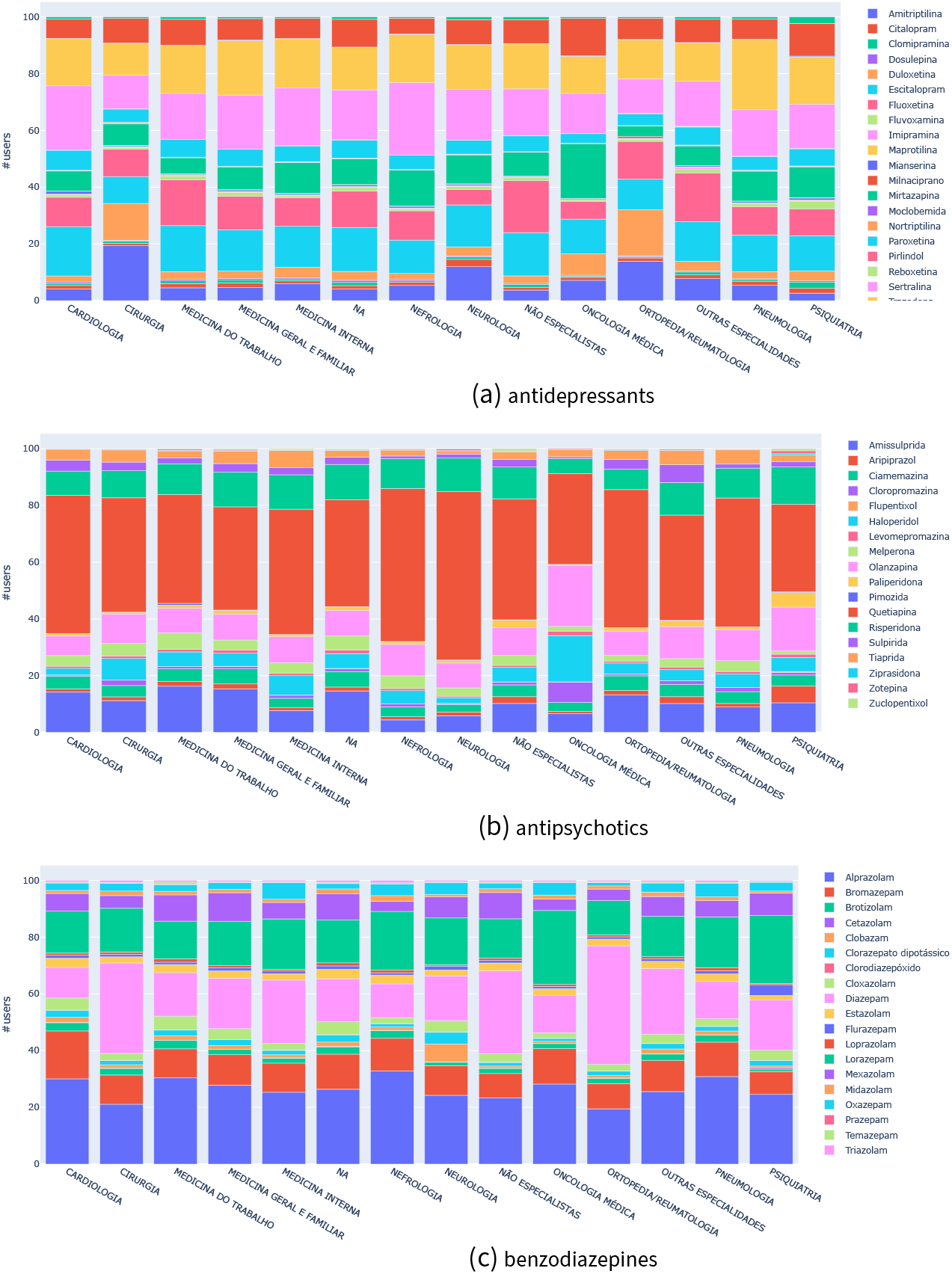
Distribution of prescribed users per psychotropic drug and medical specialty, 2018–2019.

#### B1 Psychotropic drugs

- **antidepressants**
  - selective serotonin reuptake inhibitors, including Citalopram, Escitalopram, Fluoxetina, Paroxetina, Sertralina, Fluvoxamina and Vortioxetina;
  - selective serotonin and norepinephrine reuptake inhibitors, inc. Milnaciprano, Venlafaxina, Duloxetina;
  - tricyclic antidepressants, including Amitriptilina, Imipramina, Maprotilina, Nortriptilina, Pirlindol, Trimipramina, Clomipramina, Dosulepina;
  - alpha-2 antagonists, including Mianserina, Mirtazapina and Trazodona; and
  - others not directly falling into one of the above classes, including Moclobemida and Reboxetina.
- **antipsychotics**
  - *atypical*, including Amissulprida, Aripiprazol, Melperona, Olanzapina, Paliperidona, Quetiapina, Risperidona, Sulpirida, Tiaprida, Ziprasidona, Zotepina, and Zuclopentixol; and
  - *typical*, including Ciamemazina, Cloropromazina, Flufenazina, Flupentixol, Haloperidol, Levomepromazina, and Pimozida.
- **benzodiazepines**: Alprazolam, Bromazepam, Brotizolam, Cetazolam, Clobazam, Dipotassium chlorazepate, Chlorodiazepoxide (with and without clidinium bromide), Cloxazolam, Diazepam, Estazolam, Flurazepam, Loprazolam, Lorazepam, Mexazolam, Midazolam, Oxazepam, Prazepam, Temazepam e Triazolam.

#### B2 Dosages

**Table B1.**
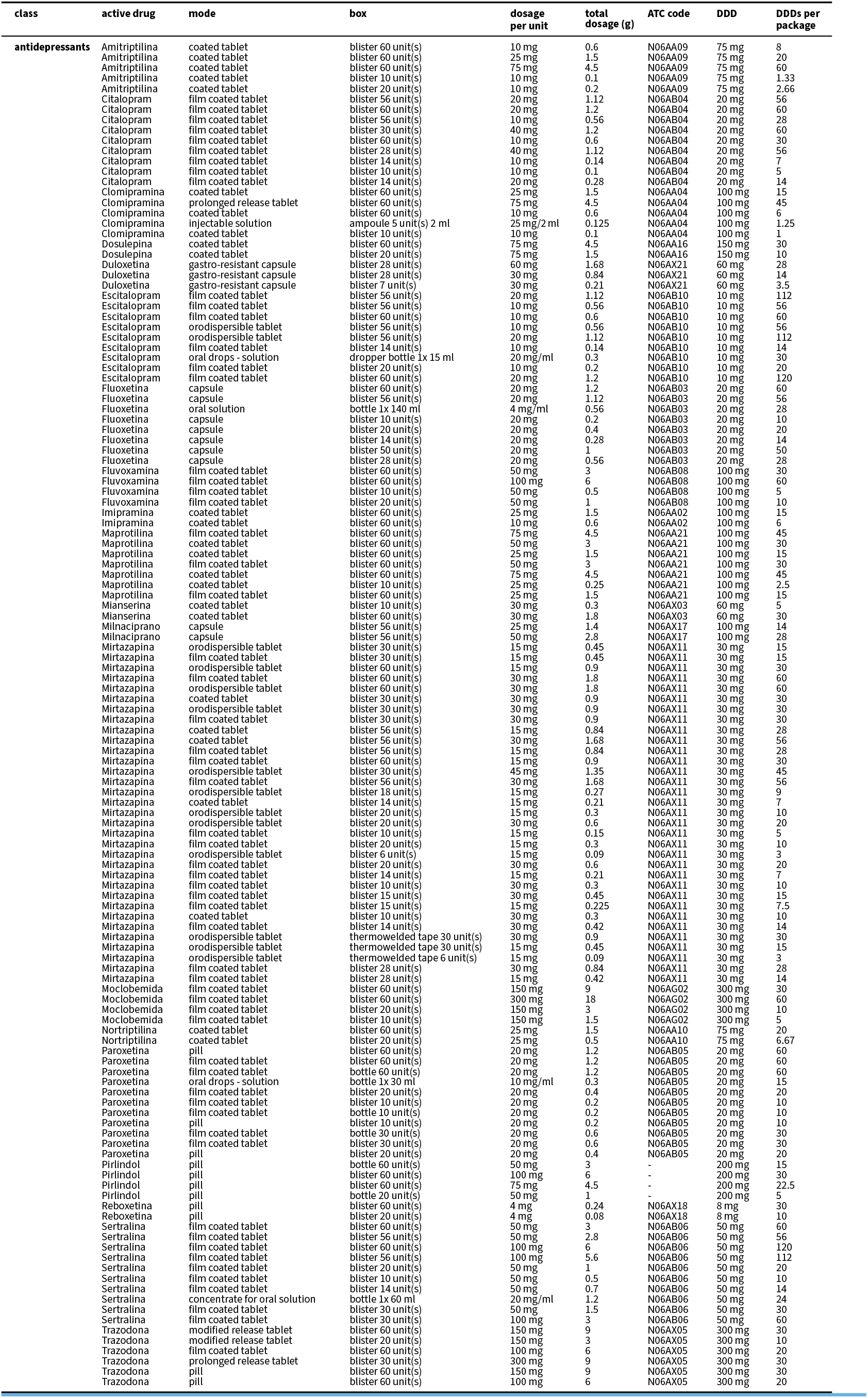
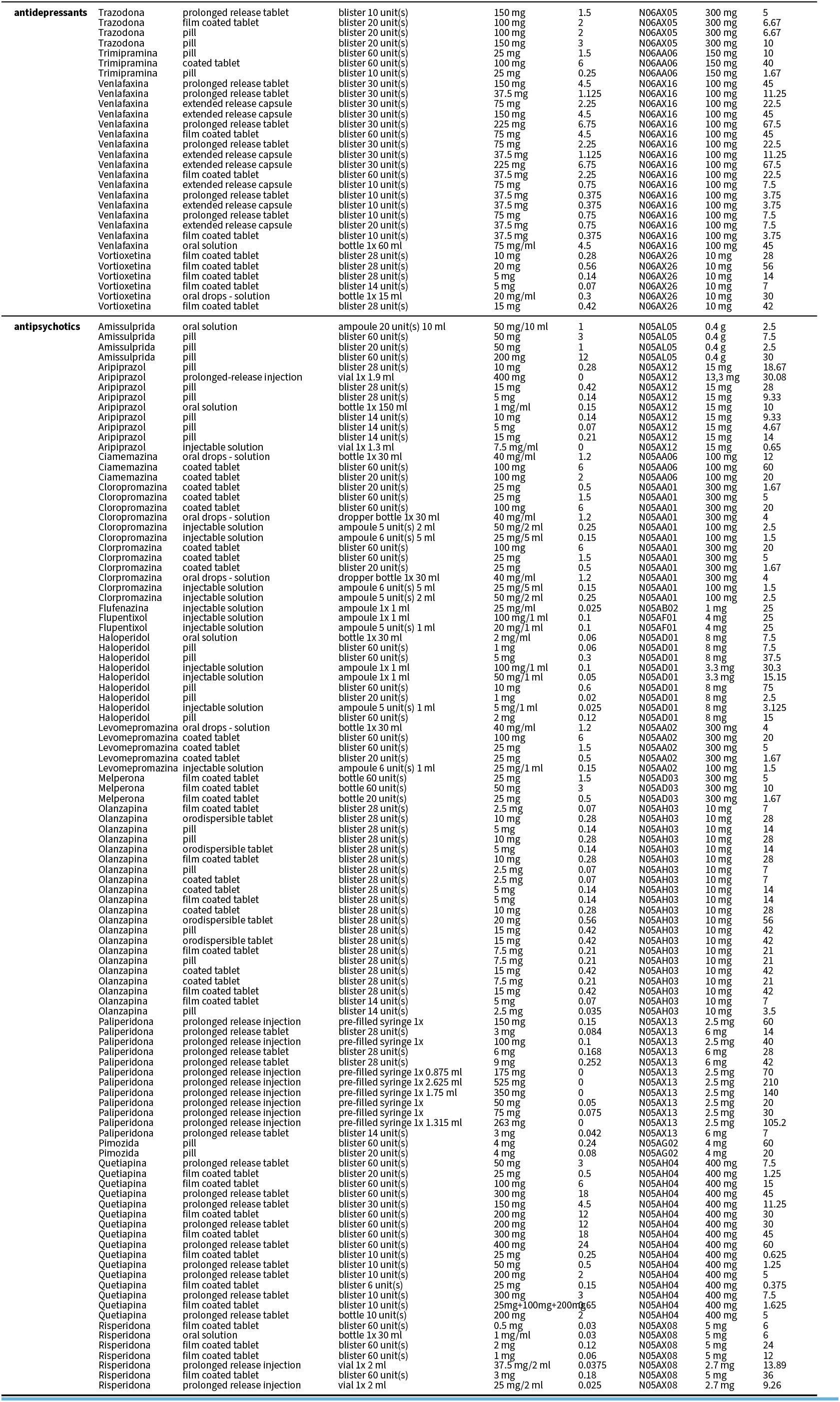
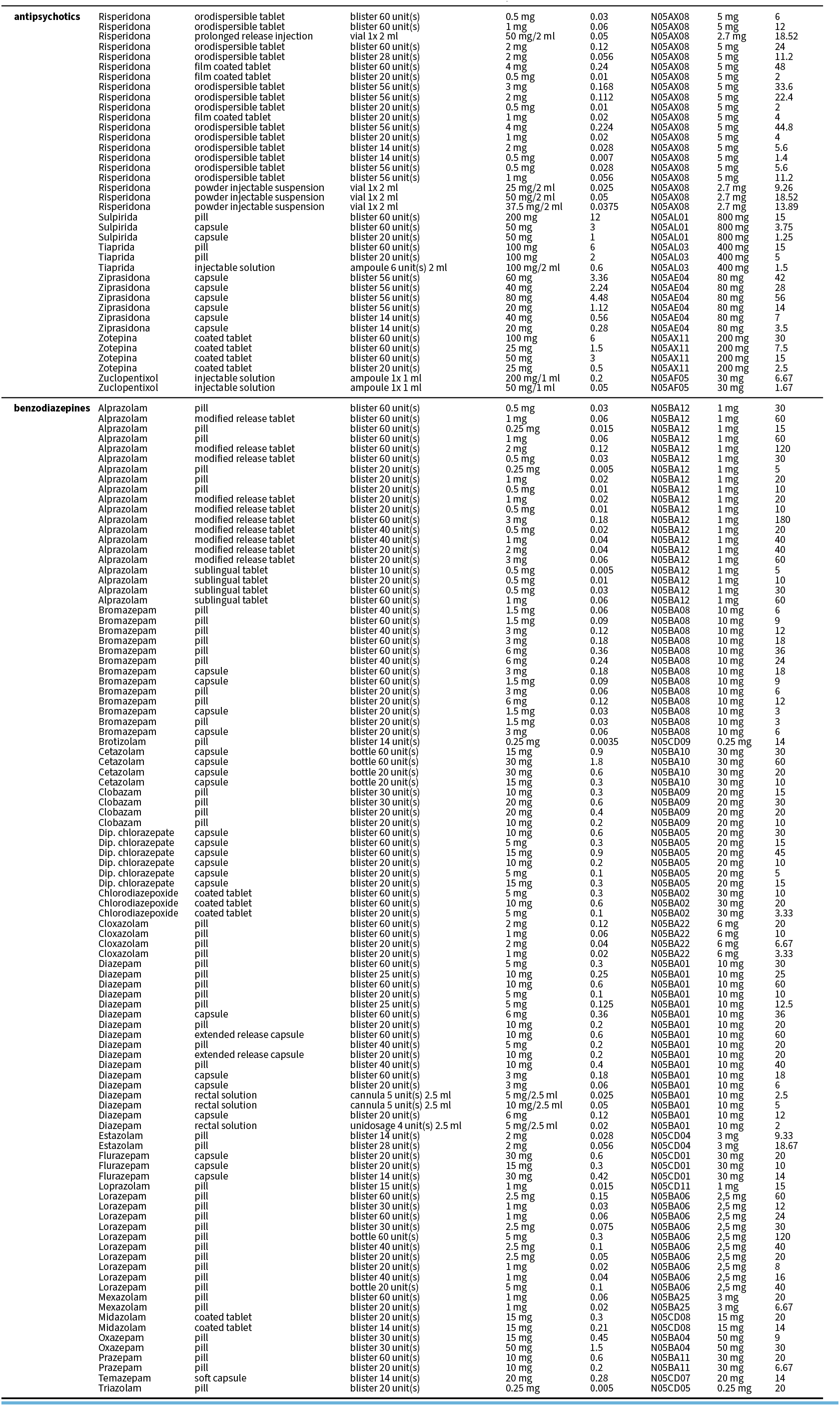
Full list of drug boxes and their corresponding ATC code, dosage, and total DDDs.

## Footnotes

1 https://www.whocc.no/atc_ddd_index/

